# Cardiac Biomarkers to Refine Pre-Test Probability for Coronary Obstruction and Predict Survival after Revascularisation in Chronic Coronary Syndrome

**DOI:** 10.64898/2025.12.18.25342624

**Authors:** Andrej Teren, Jeffrey Netto, Joachim Thiery, Holger Thiele, Christoph Stellbrink, Dennis Lawin, Thorsten Lawrenz, Anselm Artur Derda, Sylvia Henger, Holger Kirsten, Markus Scholz, Thorsten Kaiser

## Abstract

**Background:** The role of cardiovascular biomarkers in detecting coronary obstruction and predicting outcomes after revascularisation in chronic coronary syndrome (CCS) remains unclear.

**Methods:** Patients undergoing coronary angiography for suspected CCS were prospectively studied (n=2,251; median follow-up 12.6 years). Obstructive coronary artery disease (CAD) was defined as ≥50% stenosis in ≥1 major epicardial vessel. High-sensitivity cardiac troponin T (hsTnT), N-terminal pro-B-type natriuretic peptide (NT-proBNP), high-sensitivity C-reactive protein (hsCRP), interleukin-6 (IL-6), and copeptin were measured. Diagnostic performance was assessed by receiver operating characteristic (ROC); survival analysis used multivariate Cox regression with biomarker × treatment interaction testing.

**Results:** Overall 888 patients (39.4%) had obstructive CAD. Only hsTnT provided meaningful diagnostic capacity (area under the curve [AUC] 0.669), comparable to risk factor-weighted clinical likelihood (RF-CL; AUC 0.663), with **incremental dignostic benefit** inversely proportional to RF-CL category (**delta AUC: very low 10.4%, low 8.0%, intermediate/high 5.0%**). NT-proBNP was the strongest universal mortality predictor across optimal medical therapy (OMT; (hazard ratio [HR] (1.488, 95%CI 1.288–1.720, p<0.001), percutaneous coronary intervention (PCI; HR 1.220, 95%CI 1.020–1.458, p=0.029), and coronary artery bypass grafting (CABG; HR 1.220, 95%CI 1.049–1.420, p=0.010). Interaction analysis validated a data-derived 150 pg/mL threshold (p=0.032): below it, mortality was comparably low irrespective of revascularisation status (HR 0.98, 95%CI 0.67–1.43, p=0.910); above it, baseline mortality risk was markedly elevated (HR 5.75, 95%CI 4.10–8.00, p<0.001) and revascularisation associated with 40% mortality reduction, though substantial residual risk persisted (HR 3.43, 95%CI 2.70–4.40, p<0.001).

**Conclusions:** HsTnT provides meaningful diagnostic value with RF-CL category-specific incremental benefit. NT-proBNP emerged as a universal prognostic marker identifying patients with distinct revascularisation-associated mortality reduction driven by differential baseline risk.

**Clinical Trial Registration:** ClinicalTrials.gov NCT00497887

**CLINICAL PERSPECTIVE:** *What Is New?:*

- High-sensitivity troponin T provides incremental diagnostic value for coronary obstruction that follows an inverse gradient, with greatest benefit in very low pre-test likelihood patients (ΔAUC 10.4%).
- NT-proBNP ≥150 pg/mL identifies a CCS subgroup with markedly elevated baseline mortality risk in whom revascularisation is associated with a 40% mortality reduction, yet substantial residual risk persists.

*What Are the Clinical Implications?:*

- hsTnT testing adds most diagnostic value when selectively applied in low pre-test likelihood patients, while elevated NT-proBNP signals the need for comprehensive medical management and long-term surveillance beyond revascularisation alone.

## INTRODUCTION

Coronary artery disease (CAD) remains a leading cause of morbidity and mortality worldwide, with chronic coronary syndrome (CCS) representing a significant clinical burden (1,2). Accurate detection of coronary obstruction and prediction of long-term outcomes remain challenging (3,4).

Current diagnostic approaches rely on clinical assessment, non-invasive imaging, and invasive coronary angiography, with inherent limitations (5). The 2024 European Society of Cardiology (ESC) guidelines introduced a revised pre-test probability (PTP) model based on risk factor – weighted clinical likelihood (RF-CL) incorporating additional risk factors (i.e. diabetes, smoking, dyslipidemia, hypertension, and family history) beyond the traditional age-sex-symptom triad (2). Direct comparison of novel biomarker-based approaches to improve the predictive value of guideline-recommended PTP models remains limited. Moreover, differences in revascularisation benefit between CCS and myocardial infarction patients highlight the need for tailored therapeutic and secondary-preventive strategies (6).

Cardiac-specific biomarkers High-sensitivity cardiac troponin T (hsTnT) and N-terminal pro-B-type natriuretic peptide (NT-proBNP) have demonstrated utility in acute myocardial infarction (MI), and to a lesser extent in CCS settings (6–8). NT-proBNP, in particular, represents a cornerstone biomarker across the entire spectrum of cardiovascular diseases, demonstrating strong diagnostic and prognostic value in heart failure patients from acute decompensation through chronic stable disease, with guideline-recommended thresholds for diagnosis and risk stratification (7). In CCS, these biomarkers may serve dual roles in both diagnostic and prognostic applications. Inflammatory biomarkers high-sensitivity C-reactive protein (hsCRP) and interleukin-6 (IL-6), and copeptin also warrant systematic investigation, as these biomarkers may offer incremental value in both diagnostic assessment and prognostic stratification of CCS patients (3,9).

Beyond stenosis detection, identifying patients most likely to benefit from revascularisation remains an unresolved challenge (10). Recent meta-analysis of 19 randomised trials demonstrated that percutaneous coronary intervention reduces cardiovascular mortality (OR 0.77, p=0.02) in CCS, yet benefit varies across subgroups, underscoring the need for biomarker-based patient selection beyond anatomic evaluation alone (11).

This study aimed to evaluate the diagnostic performance of five circulating biomarkers against 2024 PTP model, assesses incremental diagnostic value stratified by baseline risk categories, and determines prognostic utility across different treatment strategies to identify biomarker thresholds predicting long-term survival after revascularisation.

## METHODS

### Data Availability Statement

The data that support the findings of this study are available from the Leipzig Research Centre for Civilization Diseases (LIFE), University of Leipzig. Restrictions apply to the availability of these data, which were used under license for this study. Data are available from the corresponding author upon reasonable request and with permission of the LIFE study steering committee, subject to institutional data sharing agreements and ethical approval.

### Study Design and Population

This analysis utilized data from the Leipzig Research Centre for Civilization Diseases - Heart study (LIFE-Heart study), a prospective longitudinal cohort examining patients with suspected CCS at the Heart Center Leipzig at Leipzig University. The study aimed to identify genetic, biochemical, and environmental markers associated with coronary and extracoronary atherosclerosis development (12). The protocol adhered to Declaration of Helsinki principles and was approved by the Ethics Committee of the Medical Faculty, University of Leipzig (registration 276-2005; ClinicalTrials.gov NCT00497887). Written informed consent was obtained from all participants. Blood samples were collected prospectively at enrollment prior to coronary angiography, with subsequent clinical follow-up for vital status and cardiovascular outcomes. Primary endpoint was all-cause mortality, ascertained through civil registry linkage, ensuring complete vital status follow-up.

We here consider a subset of LIFE-Heart comprising 3,305 patients (2,122 males, 1,183 females) with suspected CCS based on clinical symptoms or positive non-invasive testing, undergoing first-time coronary angiography. Exclusion criteria for the present analysis included acute myocardial infarction, acute decompensated heart failure, pregnancy, lactation, and severe systemic diseases (autoimmune diseases with immunomodulatory therapy, dialysis-requiring conditions, infectious diseases, and malignancy or cancer treatment within two years).

After excluding further 1,054 patients (821 for incomplete clinical symptoms and cardiovascular risk factor assessment, 121 for indeterminate angiographic findings, 112 for hsTnT ≥52 pg/mL), 2,251 patients were eligible for the present analysis. Median follow-up was 12.6 years (range 0.1-16.2 years).

### Assessment of Cardiovascular Risk Factors and Coronary Status

Traditional atherosclerotic risk factors were evaluated through standardized clinical interviews, biometric assessments, and laboratory analyses. Key comorbidities considered as confounding variables included diabetes mellitus (i.e. history of diabetes mellitus, or HbA1c ≥ 6.5%), obesity (i.e. BMI ≥ 30 kg/m²), dyslipidemia (i.e. history of dyslipidemia or administration of lipid-modifying agents – ATC group C10), hypertension (i.e. history of arterial hypertension, administration of antihypertensives, or current blood pressure ≥ 140/90 mmHg), and impaired renal function i.e. estimated glomerular filtration rate (eGFR) applying CKD-EPI formula < 90 mL/min/1.73 m². Medications were recorded according to Anatomical Therapeutic Chemical (ATC) classification codes.

Coronary angiography was performed following institutional protocols. Participants were stratified based on angiographic findings considering ≥50% luminal reduction as significant coronary obstruction. In order to refine the adjustment for angiographic extent of CAD, number of vessels with coronary obstruction and Gensini score was calculated using predefined standards (13). The selection of coronary therapeutic modality in case of relevant coronary obstruction, i.e. Optimal Medical Therapy (OMT), Percutaneous Coronary Intervention (PCI), or CABG (Coronary Artery Bypass Grafting), was determined at clinical discretion of the physician performing the index coronary angiography.

### Laboratory Procedures

Venous blood samples were collected at the day of index coronary angiography and transported at 4°C to the laboratory within five hours. Samples underwent immediate centrifugation, analysis, and storage in liquid nitrogen gas phase at -80°C. All procedures followed ISO 15189 and ISO 17025 standards. In the majority of patients (87.3%), samples were obtained immediately prior to catheterization (median 1.2 hours before arterial access, IQR 0.5-4.1 hours). In a subset of cases (12.7%), samples were collected shortly after angiography (median 2.3 hours post-procedure).

Standard lipid profile i.e. low-density lipoprotein cholesterol (LDL-C), high-density lipoprotein cholesterol (HDL-C), and triglycerides (TG) and plus serum creatinine were analyzed according to LIFE-Heart protocol (12). Five biomarkers were selected for comprehensive evaluation based on their established pathophysiological relevance and clinical utility in cardiovascular disease: cardiac-specific markers (hsTnT, NT-proBNP) representing myocardial injury and wall stress, inflammatory markers (hsCRP, IL-6) reflecting atherosclerotic disease activity, and copeptin as a marker of chronic neurohumoral activation. HsTnT, NT-proBNP, hsCRP, and IL-6 were measured by electrochemiluminescence on a Roche Cobas 8000 analyzer (Roche Diagnostics, Mannheim, Germany). Serum copeptin was determined using immunological trace technology (ThermoFisher/Brahms, Waltham, MA, USA).

To ensure that biomarker measurements reflected chronic cardiovascular disease states rather than acute clinical conditions, we implemented predefined upper threshold exclusions. Patients with NT-proBNP ≥1000 pg/mL were excluded to eliminate those with acute decompensated heart failure, as this threshold represents the established diagnostic cutoff for acute heart failure in the ESC guidelines (7). Copeptin values ≥13.8 pmol/L were excluded to eliminate potential cases of diabetes insipidus or acute severe illness, as this threshold approximates diagnostic cutoffs for posterior pituitary dysfunction (14). For inflammatory markers, we excluded patients with hsCRP ≥10 mg/L and IL-6 ≥43.5 pg/mL to eliminate acute inflammatory states, infections, or acute phase reactions that would confound assessment of chronic cardiovascular inflammation. Importantly, for biomarkers other than hsTnT, exclusions were applied only when examining the diagnostic or prognostic value of the corresponding biomarker.

### Statistical Analysis

Statistical analyses were performed using SPSS version 29.0 and R statistical software. Continuous variables are presented as medians with interquartile ranges; categorical variables as frequencies and percentages. Complete case analysis was performed. Among 2,251 patients in the primary cohort, missing data were minimal: waist-hip ratio 93 patients (4.1%), ejection fraction 34 patients (1.5%), follow-up time 6 patients (0.3%). Patients with missing outcome or covariate data were excluded from respective analyses (sensitivity analysis cohort: 718/766 = 93.8% retention). Complete missing data analysis is provided in Supplementary Table S6. Missing data did not substantially differ by CAD status or mortality. Patients were dichotomized based on angiographic findings: obstructive CAD (i.e., ≥50% stenosis in at least one major epicardial vessel) versus no or non-obstructive disease (i.e., <50% stenosis).

Pre-test probability was assessed using RF-CL (i.e. age, sex, symptoms, diabetes, current smoking, dyslipidemia, hypertension, family history of premature CAD defined as MI or coronary revascularisation before age 60 years in first-degree relative). In order to examine the incremental discrimination value of biomarkers patients were subsequently categorized based on RF-CL as: Category 1 – very low likelihood (PTP <5%), Category 2 – low likelihood (5-15%), Category 3 – intermediate / high likelihood (>15%) of relevant coronary obstruction. The more comprehensive cardiovascular risk factor (CRF) model included RF-CL variables plus LDL-C, HDL-C, TG, body mass index (BMI), waist-hip ratio (WHR), administration of antihypertensive medication, administration of statins, history of hyperlipoproteinemia, systolic and diastolic blood pressure (averaged from 3 standardised measurements) left ventricular ejection fraction (EF) assessed by standardised transthoracic echocardiography and estimated glomerular filtration rate (eGFR) applying CKD-EPI-Formel.

For diagnostic performance assessment, Receiver Operating Characteristic (ROC) curves were constructed for all five biomarkers. Area Under the Curve (AUC) values with 95% confidence intervals were tested for differences using DeLong’s method (15). The biomarker demonstrating meaningful diagnostic performance was then compared directly with RF-CL model. Optimal cutoff was determined using Youden’s index (16). **Incremental discriminatory value was quantified by the change in AUC (delta AUC) obtained by adding each biomarker at its optimal cutoff to the baseline model, assessed both overall and within predefined RF-CL categories.** Bootstrap validation with 500 iterations established robust confidence intervals (8). Bootstrap internal validation (250 iterations) assessed optimistic bias for data-derived thresholds following Harrell-Steyerberg methodology, re-deriving optimal cutoffs in each bootstrap sample and calculating optimism as the mean difference between apparent and test performance. Model calibration was evaluated using Hosmer-Lemeshow goodness-of-fit test and calibration plots comparing observed versus predicted probabilities across deciles of predicted risk. Brier score assessed overall model performance, with lower scores indicating better calibration. Clinical utility of identified biomarker diagnostic value was evaluated through decision curve analysis (DCA), quantifying net benefit across threshold probabilities by comparing true positives gained against false positives incurred.

Survival analysis employed Kaplan-Meier curves with log-rank tests and Cox proportional hazards regression to perform safety analysis for false negatives to detect coronary obstruction (i.e. CAD cases reclassified downward based on biomarker profile) and to determine which biomarker provided strongest prognostic discrimination. Patients were stratified by coronary anatomy and treatment strategy, i.e. obstructive CAD patients treated OMT alone vs with PCI, and CABG, respectively.

Cox proportional hazards regression with backward stepwise elimination was performed using differential adjustment strategies: safety analyses for discriminative value to detect coronary obstruction adjusted for RF-CL variables, while prognostic models adjusted for number of vessels with coronary obstruction, Gensini score, and comprehensive cardiovascular risk factors (CRF). Hazard ratios were calculated per 1 SD increase to enable cross-biomarker comparison. To assess the stability of biomarker effect estimates, sensitivity analyses were performed forcing all candidate covariates into logistic and cox regression models without selection (ENTER method) (Supplementary Table S8A and S8B). Proportional hazards assumptions were verified using Schoenfeld residuals and log-log plots for all Cox models. No substantial violations were detected for primary covariates (all p>0.05 for time-dependence tests; (Supplementary Figure S1).

Time-dependent ROC analysis using the survivalROC package in R determined optimal NT-proBNP cutoff for mortality prediction. Cumulative sensitivity and specificity were calculated across biomarker values at 1, 3, 5 and 10 years, with optimal cutoff identified using Youden’s index. Bootstrap validation (500 iterations) established confidence intervals. Bootstrap validation (250 iterations) was performed to assess threshold optimism, re-deriving optimal cutoffs in each bootstrap sample using time-dependent ROC analysis for 5-year mortality.

To rigorously address potential confounding by indication, we performed 1:1 propensity score matching (PSM) without replacement using all known confounders for the subsequent analysis. Propensity scores for receiving revascularisation were estimated using multivariable logistic regression. Treated patients were matched to controls using nearest-neighbor matching with caliper width set at 0.2 times the standard deviation of the logit propensity score. Matching quality was assessed using standardized mean differences (SMD), with |SMD| <0.1 indicating excellent balance and |SMD| <0.2 considered acceptable. Covariate balance is detailed in Supplementary Table S7 and visualized in Supplementary Figure S2.

In the matched cohort, we performed Cox proportional hazards regression with minimal covariate adjustment (age, sex) given the achieved balance. The NT-proBNP × revascularisation interaction was analyzed using a four-group framework with ’No Revascularisation + NT-proBNP below the post-hoc data-derived threshold’ as the reference group, and tested using a product term interaction. Additionally, a continuous dose-response modeling was performed using restricted cubic splines.

All statistical tests were two-sided, and p-values <0.05 were considered statistically significant. **Given the exploratory and hypothesis-generating nature of this single-centre observational study, no formal correction for multiple comparisons was pre-specified. All five biomarkers were evaluated systematically for both diagnostic and prognostic utility; no single analysis was designated confirmatory a priori. All p-values are reported continuously, and all findings should be interpreted as hypothesis-generating.**

## RESULTS

### Baseline Characteristics

Among 2,251 patients, 888 (39.4%) had obstructive CAD. Table 1 presents baseline characteristics stratified by angiographic findings. Patients with obstructive CAD were older (median 67 versus 63 years, p<0.001), more frequently male (75.7% versus 60.9%, p<0.001), and exhibited higher prevalence of traditional cardiovascular risk factors.

**Table 1.** Baseline Characteristics of Study Population Stratified by Presence of Obstructive CCS.

| Characteristics | No CAD<br>N / Median | % / 25-75<br>IQR | CAD<br>N / Median | % / 25-75<br>IQR | p-value |
| --- | --- | --- | --- | --- | --- |
| **N (%)** | 1363 | 60.6% | 888 | 39.4% | - |
| **Female sex (N)** | 605 | 44.4% | 193 | 21.7% | < 0.001 |
| **Age (years)** | 61.6 | 53.8-69.8 | 65.7 | 57.3-71.1 | < .001 |
| **BMI (kg/m<sup>2</sup>)** | 29.3 | 26.4-32.8 | 29.2 | 26.4-32.6 | .370 |
| **Waist-hip ratio** | 0.96 | 0.88-1.03 | 1.01 | 0.95-1.01 | < .001 |
| **Diabetes (N)** | 335 | 24.6% | 302 | 34.0% | < .001 |
| **Family history with MI (N)** | 158 | 11.6% | 113 | 12.7% | .439 |
| **Current smoker (N)** | 213 | 15.6% | 188 | 21.2% | .003 |
| **Systolic blood pressure (mmHg)** | 136 | 125-149 | 138 | 126-153 | .095 |
| **Diastolic blood pressure (mmHg)** | 84.5 | 77.5-92.0 | 82.5 | 74.6-89.4 | < .001 |
| **Antihypertensive medication (N)** | 1144 | 83.9% | 776 | 87.4% | < .001 |
| **Hyperlipidemia (N)** | 1165 | 85.5% | 817 | 92.0% | .002 |
| **Lipid lowering medication (N)** | 459 | 33.7% | 417 | 47.0% | < .001 |
| **LDL-cholesterol (mmol/l)** | 3.26 | 2.61-3.91 | 3.43 | 2.66-4.23 | < .001 |
| **HDL-cholesterol (mmol/l)** | 132 | 1.10-1.61 | 1.22 | 1.01-1.48 | < .001 |
| **Triglycerides (mmol/l)** | 1.63 | 1.13-2.41 | 1.74 | 1.27-2.48 | < .001 |
| **eGFR (in ml/min/1.73 m<sup>2</sup>)** | 87.8 | 75.2-96.8 | 84.9 | 71.9-94.8 | < .001 |
| **Left-ventricular ejection fraction (%)** | 61 | 55-66 | 60 | 54-65 | .022 |
| **Typical Angina** | 466 | 34.2% | 437 | 49.2% | < .001 |
| **Atypical Angina** | 213 | 15.6% | 85 | 9.6% | < .001 |
| **Non-anginal Chestpain** | 316 | 23.2% | 109 | 12.3% | - |
| **Dyspnoea NYHA $\geq 2$** | 709 | 52.0% | 474 | 53.4% | .546 |
| **No chest pain / no Dyspnoe** | 262 | 11.6% | 193 | 8.6% | .407 |
| **ESC PTP 2024 5%** | 260 | 19.1 | 47 | 5.3 | < .001 |
| **5% - 15%** | 637 | 46.7 | 313 | 35.2 |  |
| **> 15%** | 466 | 34.2 | 528 | 59.2 |  |
| **PCI** | - | - | 344 | 38.7 | - |
| **CABG** | - | - | 267 | 30.1 |  |
| **hsTnT (pg/ml)** | 6.58 | 4.0-10.7 | 9.86 | 6.55-15.56 | < .001 |
| **NT-pro BNP (pg/ml)** | 111.5 | 50.6-272.2 | 172.1 | 75.3-415.7 | < .001 |

| Characteristics | No CAD | % / 25-75 | CAD | % / 25-75 | p-value |
| --- | --- | --- | --- | --- | --- |
|  | N / Median | IQR | N / Median | IQR |  |
| **CRP (mg/l)** | 1.96 | 1.02-4.19 | 2.35 | 1.12-5.07 | < .001 |
| **IL-6 (pg/ml)** | 2.38 | 1.50-4.02 | 2.99 | 1.80-5.50 | < .001 |
| **Copeptin (pmol/l)** | 4.72 | 3.05-7.86 | 5.91 | 3.86-9.85 | < .001 |

*Abbreviations: CAD, obstructive coronary artery disease; BMI, body mass index; MI, myocardial infarction; LDL, low-density lipoprotein; HDL, high-density lipoprotein; eGFR, estimated glomerular filtration rate; NYHA, New York Heart Association; ESC, European Society of Cardiology; PTP, pre-test probability; PCI, percutaneous coronary intervention; CABG, coronary artery bypass grafting; hsTnT, high-sensitivity Troponin T; NT-proBNP, N-terminal pro-B-type natriuretic peptide; CRP, C-reactive protein; IL-6, Interleukin 6; IQR, interquartile range.*

Left ventricular function was preserved (EF ≥50%) in 1,850 patients (82.2%), with mildly reduced (40-49%) in 267 (11.9%) and reduced (<40%) in 100 (4.4%). Heart failure symptom burden by NYHA functional class ≥2 was documented in 1183 patients (52.6%).

Biomarker concentrations demonstrated systematic elevation in patients with obstructive CAD. Median hsTnT was 10.0 ng/L (IQR 6.0-16.0) in obstructive CAD versus 7.0 ng/L (IQR 5.0-11.0) in non-obstructive disease (p<0.001). NT-proBNP showed similar gradient: 151 pg/mL (IQR 74-340) versus 98 pg/mL (IQR 49-198), p<0.001. Inflammatory markers exhibited more modest differences.

Obstructive CAD prevalence showed progressive separation across RF-CL categories (low-likelihood 23.6%, intermediate 32.9%, high-likelihood 53.1%) (chi-square p<0.001).

### Diagnostic Performance of Biomarkers to detect anatomical coronary obstruction

Comparative ROC analysis across all five biomarkers revealed marked heterogeneity in diagnostic performance for detecting anatomical coronary obstruction. Only hsTnT demonstrated clinically relevant diagnostic capacity with AUC 0.669 (95% CI: 0.644-0.692). NT-proBNP showed modest discrimination with AUC 0.609 (95% CI: 0.585-0.633), whilst inflammatory markers demonstrated limited diagnostic value: hsCRP AUC 0.536 (95% CI: 0.511-0.561), IL-6 AUC 0.547 (95% CI: 0.522-0.572), and copeptin AUC 0.524 (95% CI: 0.499-0.549).

Direct comparison with ESC 2024 PTP model (Figure 1) demonstrated that hsTnT performance was comparable to RF-CL (AUC 0.663, 95% CI: 0.642-0.689, p=0.842 for comparison). The comprehensive CRF model achieved strong baseline performance (AUC 0.731, 95% CI: 0.709-0.753), with hsTnT addition providing limited additional benefit (AUC 0.747, improvement +0.016, p = 0.013). Optimal hsTnT cutoff by Youden index was 7.0 ng/L (sensitivity 72.4%, specificity 54.0%) (Supplementary Table S1). This threshold provided the basis for incremental value assessment beyond current RF-CL model (Supplementary Table S1). Bootstrap validation (250 iterations) confirmed minimal bias: optimal cutoff median 6.97 ng/L (95% CI: 6.62-8.47), chosen 7.0 ng/L at 70th percentile, AUC optimism 0.001 (optimism-corrected AUC 0.668).

**Figure 1:**
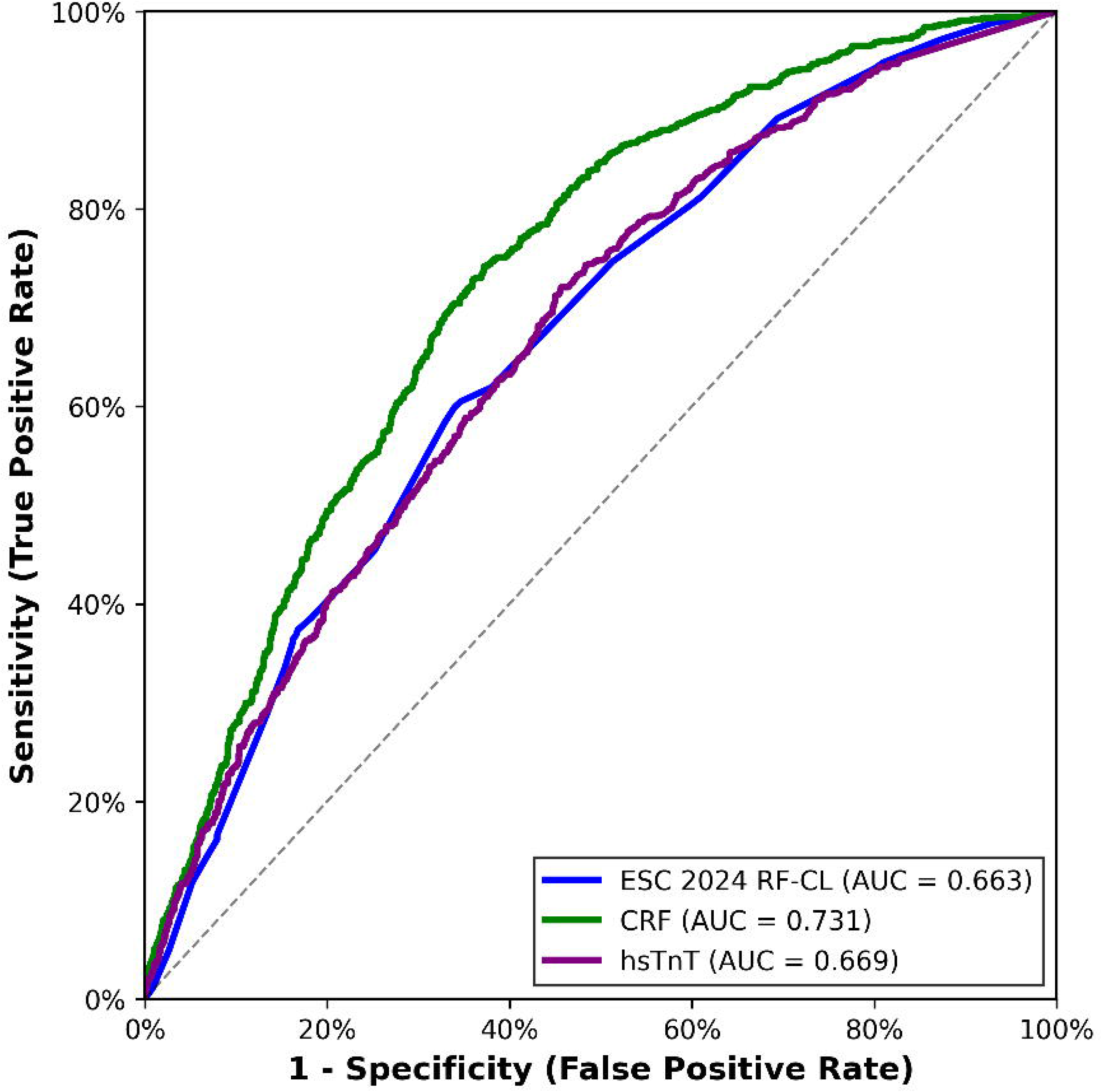
Comparative Diagnostic Performance of hsTnT with ESC 2024 PTP. (considering age, sex, symptoms plus diabetes, smoking, dyslipidemia, hypertension, family history of premature CAD). *Abbreviations: CAD, coronary artery disease, hsTnT, high-sensitivity Troponin T; ESC, European Society of Cardiology; PTP, pre-test probability; CRF, comprehensive cardiovascular risk factor model (i.e. ESC 2024 PTP variables plus LDL, low-density lipoprotein; HDL, high-density lipoprotein; TG, triglycdrides; BMI, body mass index; WHR, waist-hip ratio; EF, left-ventricular ejection fraction; eGFR, estimated glomerular filtration rate)*.

Incremental value of the hsTnT 7.0 ng/L cutoff stratified by RF-CL categories demonstrated an inverse relationship between baseline pre-test clinical likelihood and biomarker benefit (Table 2, Supplementary Table S1). In very low likelihood patients (RF-CL <5%), hsTnT addition yielded substantial **discrimination improvement (delta AUC = 10.4%, p<0.001)**. Low pre-test likelihood patients (RF-CL 5-15%) showed moderate benefit: **delta AUC =8.0%, p<0.001**. Intermediate/high likelihood patients (RF-CL >15%) exhibited the smallest incremental value: **delta AUC =5.0%, p<0.001** (Table 2).

**Table 2.** AUC Improvement by ESC 2024 RF-CL Risk Category to detect coronary obstruction. Comparative discriminative ability of RF-CL model alone versus combined with hsTnT, stratified by baseline risk categories.

| **RF-CL Category** | **N** | **Prevalence**<br>**obstructive**<br>**CAD (%)** | **AUC**<br>**(RF-CL)** | **AUC (RF-**<br>**CL+ hsTnT**<br>**7.0 ng/L)** | **Delta AUC 95%**<br>**CI (%)** | **p**<br>**value** |
| --- | --- | --- | --- | --- | --- | --- |
| Very Low (<5%) | 307 | 15.3% | .6 | .704 | .104 (.062-.146) | < .001 |
| Low (5-15%) | 950 | 32.9% | .531 | .611 | .08 (.052-.108) | < .001 |
| Intermediate/High (>15%) | 994 | 53.1% | .543 | .593 | .05 (.024-.076) | < .001 |
| Overall | 2,251 | 39.4% | .663 | .694 | .031 (-.001-.064) | < .001 |

*Abbreviations: CAD, coronary artery disease, ESC, European Society of Cardiology; RF-CL, risk factor weighted clinical likelihood; hsTnT, high-sensitivity Troponin T; AUC, area under the curve.*

Model calibration analysis (Supplementary Figure S3A) demonstrated that the RF-CL model showed acceptable fit (Hosmer-Lemeshow χ²=12.4, p=0.134, Brier score=0.224), with addition of hsTnT ≥7.0 ng/L cutoff improving calibration (χ²=8.9, p=0.351, Brier score=0.216, ΔBrier=-0.008; p<0.001). Decision curve analysis (Supplementary Figure S3B) revealed that hsTnT provided clinical net benefit across threshold probabilities of 20-60%, with greatest benefit in the 30-45% range.

Reclassification patterns (Supplementary Table S2) showed differential biomarker impact by disease status: among 888 patients with obstructive CAD, 218 (24.5%) were reclassified upward and 228 (25.7%) downward, while among 1,363 patients without obstructive CAD, 521 (38.2%) were appropriately reclassified downward versus 312 (22.9%) upward. Safety analysis (Supplementary Table S3) revealed that unadjusted analysis showed false negative patients (obstructive CAD with hsTnT <7.0 ng/L, n=249) had significantly higher mortality than true negatives (no obstructive CAD with hsTnT <7.0 ng/L) in the very low likelihood category (HR=1.83, 95% CI: 1.16-2.89, p=0.009). However, adjustment for RF-CL risk factor variables substantially attenuated this apparent excess mortality to non-significance: very low CL (adjusted HR=1.29, 95% CI: 0.81-2.06, p=0.283), low CL (adjusted HR=0.90, 95% CI: 0.53-1.54, p=0.707), and overall (adjusted HR=1.10, 95% CI: 0.77-1.56, p=0.608). RF-CL factors as such (i.e. not used as scoring components in categorical RF-CL model) explained 65-81% of the excess risk, demonstrating that the unadjusted mortality differences were primarily attributable to baseline cardiovascular risk factor burden rather than low hsTnT itself.

### Long-Term Survival Outcomes

During median 12.6-year follow-up, 684 deaths occurred (30.4%) in 2,251 patients. Overall survival rates were: no/non-obstructive CAD 74% (95% CI: 72-77%, n=1,363), obstructive CAD with PCI 69% (64-74%, n=344), OMT 64% (58-69%, n=277), and CABG 54% (48-60%, n=267; log-rank p<0.001). No/non-obstructive CAD patients had significantly better survival than all obstructive CAD groups (all p≤0.016), PCI showed superior survival versus both OMT (p=0.038) and CABG (p<0.001), while OMT and CABG did not differ significantly (p=0.16). Figure 2 presents full-adjusted Cox regression hazard ratios across patient subgroups. NT-proBNP demonstrated universal prognostic utility across all categories with consistent effect magnitudes. Per 1 SD increase, NT-proBNP conferred increased mortality risk: OMT HR 1.488 (95% CI: 1.288-1.720, p<0.001), PCI HR 1.220 (95% CI: 1.020-1.458, p=0.029), CABG HR 1.220 (95% CI: 1.049-1.420, p=0.010).

**Figure 2:**
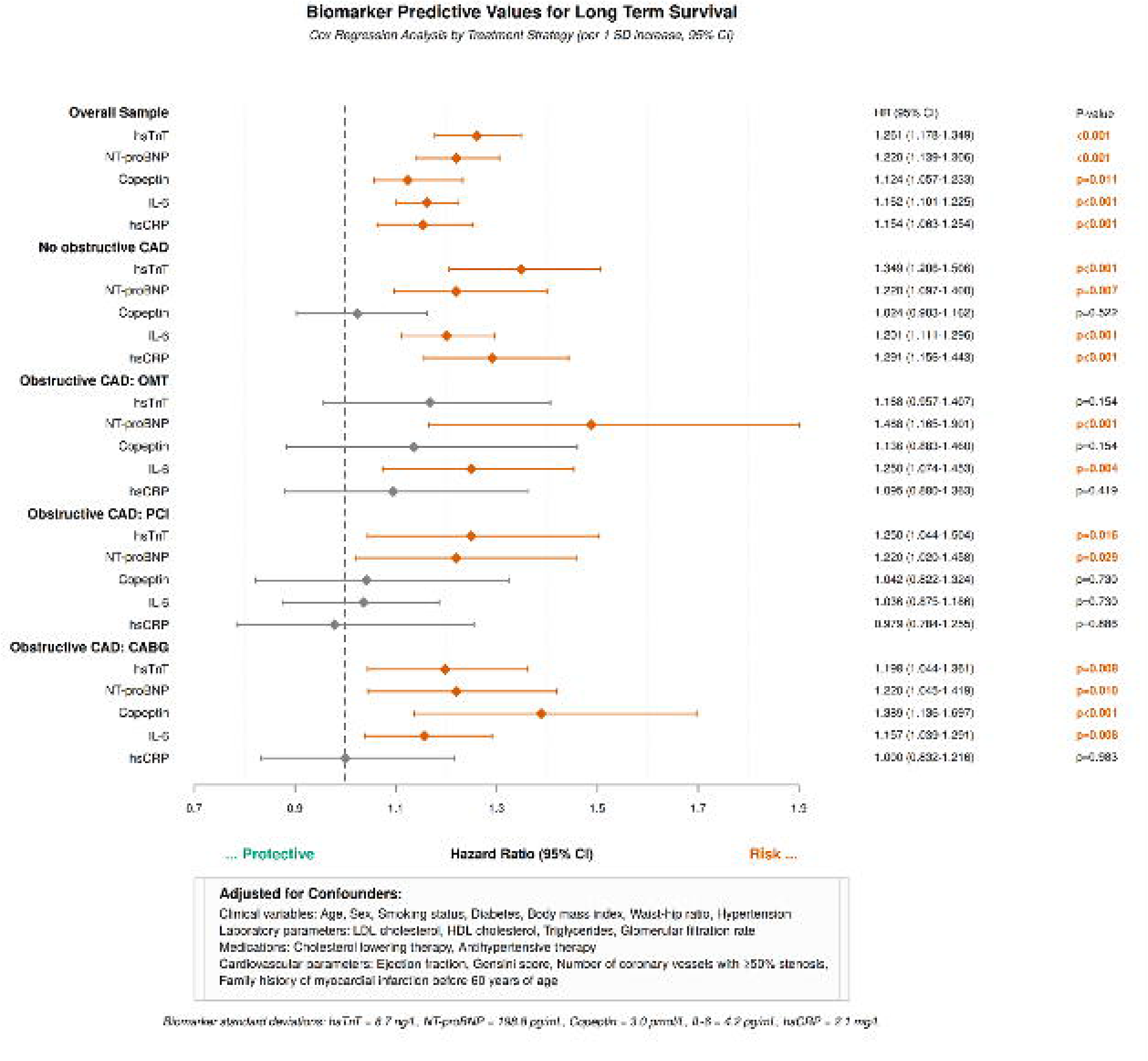
Treatment-Stratified Biomarker Prognostic Performance. Cox regression hazard ratios (per 1 SD increase) for five biomarkers across patient subgroups. Significant associations (p<0.05) shown in red (increased risk); non-significant in gray. NT-proBNP demonstrated universal prognostic utility across all categories. *Abbreviations: SD, standard deviation; CAD, coronary artery disease; hsTnT, high-sensitivity Troponin T; NT-proBNP, N-terminal pro-B-type natriuretic peptide; CRP, C-reactive protein; IL-6, Interleukin 6; LDL, low-density lipoprotein; HDL, high-density lipoprotein; OMT, optimal medical therapy; PCI, percutaneous coronary intervention; CABG, coronary artery bypass grafting*.

Other biomarkers showed inconsistent prognostic associations. HsTnT and hsCRP maintained significance in OMT and non-obstructive CAD but not in revascularised patients. Notably, copeptin demonstrated the strongest prognostic association in CABG patients (HR 1.389, 95% CI: 1.130-1.697, p=0.001), exhibiting the highest hazard ratio among all biomarkers in this treatment subgroup.

Time-dependent ROC analysis across multiple follow-up time points (1, 3, 5, and 10 years) was performed separately for PCI (n=344, 108 deaths) and CABG (n=267, 99 deaths) subgroups using Youden’s Index optimization. Optimal NT-proBNP cutoffs varied across time points: PCI subgroup ranged from 119.4 pg/mL (5-year) to 233.1 pg/mL (1-year); CABG subgroup ranged from 142.8 pg/mL (5-year) to 175.9 pg/mL (1-year). Based on this analysis, we propose 150 pg/mL as a threshold, approximating the median across all time points and both revascularisation strategies (median 153.4 pg/mL; range 119.4-233.1 pg/mL) while aligning with long-term mortality prediction cutoffs most relevant for sustained treatment effects. Among all biomarkers evaluated, NT-proBNP demonstrated the most consistent discriminative performance across intervention types, with excellent prognostic capacity at 5 years post-revascularisation. Patients with NT-proBNP <150 pg/mL (n=1,219) achieved 10-year survival of 90.6% versus 68.8% in those ≥150 pg/mL (n=1,026; fully adjusted HR 1.69, 95% CI: 1.41-2.03, p<0.001) in fully adjusted model including coronary disease severity and ejection fraction (Figure 3). Bootstrap validation (250 iterations) confirmed threshold robustness: optimal cutoff median 128.8 pg/mL (95% CI: 119.2-199.0), chosen 150 pg/mL at 74th percentile, negligible AUC optimism (-0.000).

**Figure 3:**
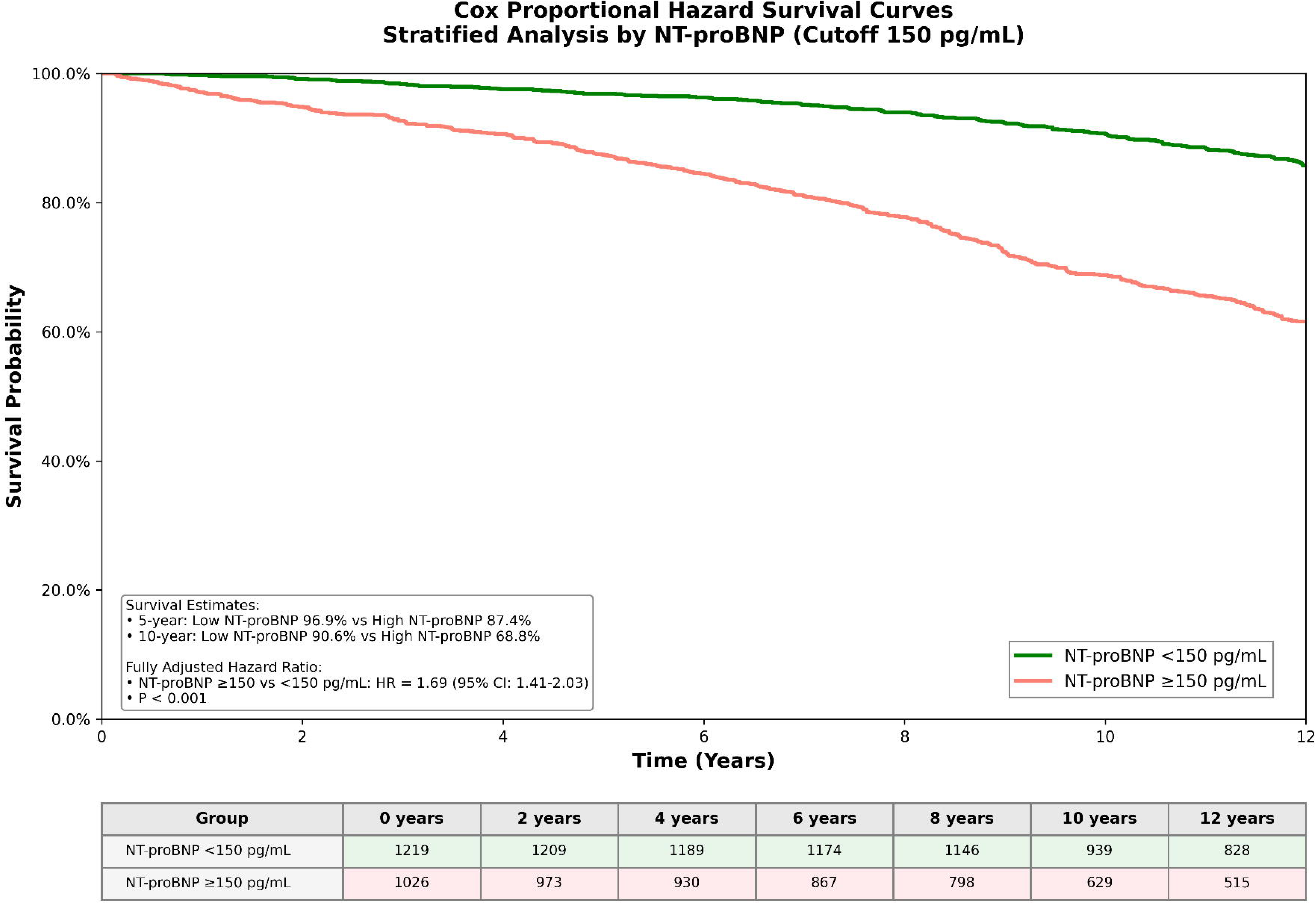
Cox Proportional Hazard Survival Curves Stratified by NT-proBNP Cutoff 150 pg/mL Cox proportional hazards survival analysis demonstrating prognostic divergence based on NT-proBNP stratification at 150 pg/mL cutoff. Green curve: NT-proBNP <150 pg/mL (n=1,219 patients); Red curve: NT-proBNP ≥150 pg/mL (n=1,026 patients). Survival probabilities: 5-year (96.9% vs 87.4%), 10-year (90.6% vs 68.8%). *Abbreviations: NT-proBNP, N-terminal pro-B-type natriuretic peptide*.

Kaplan-Meier 10-year survival estimates by NT-proBNP level and treatment strategy revealed that among patients with NT-proBNP <150 pg/mL, OMT group achieved 91.2% survival versus 90.8% in revascularisation group (absolute risk difference +0.4%, 95% CI: -3.2% to +4.0%). Among patients with NT-proBNP ≥150 pg/mL, OMT group achieved 68.1% survival versus 76.4% in revascularisation group (absolute risk difference +8.3%, 95% CI: +3.1% to +13.5%).

For the subsequent analysis, PSM yielded 572 matched pairs (n=1,144 total) comparing revascularisation vs. OMT. Excellent covariate balance was achieved (mean absolute standardized mean difference 0.069, range 0.001-0.142; Supplementary Table S7, Supplementary Figure S2), with all covariates showing |SMD|<0.15.

Using a four-group framework with no revascularisation + NT-proBNP <150 pg/mL as reference with the lowest risk, we observed distinct mortality patterns (Figure 4). In patients with NT-proBNP <150 pg/mL revascularisation associated with mortality comparable to the low risk reference subgroup (HR 0.98, 95% CI 0.67-1.43, p=0.910). Patients with NT-proBNP ≥150 pg/mL demonstrated markedly elevated baseline risk (HR 5.75, 95% CI 4.10-8.00, p<0.001 without revascularisation) with 40% relative mortality reduction following revascularisation (HR 3.43, 95% CI 2.70-4.40, p<0.001), though substantial residual risk persisted. The interaction term confirmed differential treatment effect (HR 0.609, 95% CI 0.387-0.959, p=0.032). Absolute mortality rates over 12.6 years were 18.1% (reference), 17.6% (low NT-proBNP + revascularisation), 52.1% (high NT-proBNP alone), and 38.4% (high NT-proBNP + revascularisation). Restricted cubic spline modelling demonstrated a non-linear trend between continuous NT-proBNP and mortality risk (p for non-linearity = 0.057), with the dose-response curve suggesting three distinct prognostic phenotypes that broadly correspond to the dichotomous threshold identified at 150 pg/mL (Supplementary Figure S4).

**Figure 4:**
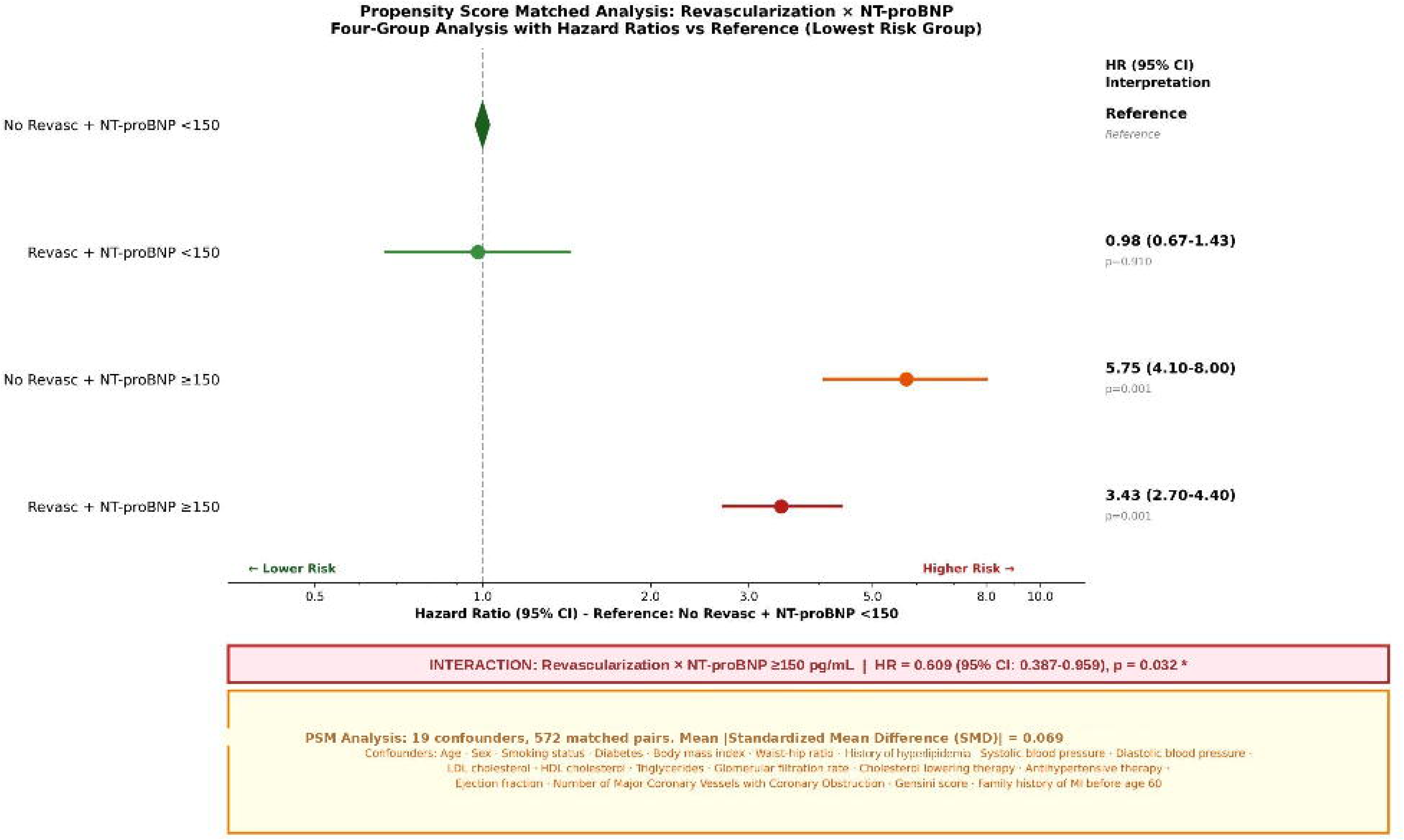
Propensity Score Matched Interaction Analysis – Revascularisation × NT-proBNP. Forest plot demonstrating significant NT-proBNP × Revascularisation interaction (interaction term HR=0.609, 95% CI: 0.387–0.959, p=0.032) from propensity score matched analysis (19 confounders, 572 matched pairs, mean |SMD|=0.069). The reference group comprises patients without revascularisation and NT-proBNP <150 pg/mL. Hazard ratios for the three remaining groups are expressed relative to this reference. *Abbreviations: HR, hazard ratio; SMD, Standardized Mean Difference; Revasc, Revascularisation i.e. PCI, percutaneous coronary intervention or CABG, coronary artery bypass grafting; NT-proBNP, N-terminal pro-B-type natriuretic peptide; LDL, low-density lipoprotein; HDL, high-density lipoprotein; OMT, optimal medical therapy*.

## DISCUSSION

This comprehensive biomarker evaluation in CCS patients yields three principal findings: (1) among five biomarkers assessed, only hsTnT provides clinically meaningful diagnostic capacity for obstructive CAD (AUC 0.669), with performance comparable to RF-CL assessment (0.663) and demonstrating inverse relationship between baseline RF-CL and incremental discriminative value; (2) NT-proBNP emerges as superior prognostic biomarker with universal discrimination across all treatment categories; (3) NT-proBNP 150 pg/mL threshold identifies distinct prognostic phenotypes with differential survival outcomes by coronary revascularisation status. These findings extend our prior work (17) through direct comparison with the currently recommended PTP model, quantification of the incremental diagnostic value of hsTnT across RF-CL categories, and identification of a NT-proBNP threshold associated with differential survival patterns by revascularisation status — addressing critical needs for biomarker-guided treatment selection in contemporary CCS management.

HsTnT’s diagnostic capacity reflects its direct relationship to myocardial injury from flow-limiting stenoses through repetitive ischemia-reperfusion (18,19), achieving performance comparable to ESC 2024 PTP. Clinical likelihood-stratified analysis with the optimized 7.0 ng/L cutoff demonstrated marked heterogeneity: patients with very low pre-test likelihood (**delta AUC=10.4%**) showed greatest benefit, moderate benefit in low likelihood patients, while intermediate/high likelihood patients (**delta AUC=5.0%)** showed the smallest benefit. This inverse gradient pattern likely reflects information content redundancy: in patients with multiple traditional risk factors and typical symptoms (high RF-CL), biomarker measurement adds limited new information, whereas in patients with few risk factors and atypical symptoms (low RF-CL), biomarker elevation meaningfully shifts probability estimates after adjustment for RF-CL variables. Crucially, safety validation demonstrated that false negative patients showed no excess mortality after adjustment for RF-CL variables, supporting the clinical applicability of this biomarker-guided approach. However, the substantial unadjusted mortality risk attributable to baseline cardiovascular risk factors underscores that safe reclassification does not obviate aggressive risk factor optimization or ongoing surveillance. A very low likelihood classification should, even after hsTnT implementation, guide deferred rather than omitted testing, with reassessment frequency individualized to baseline risk burden (1,2,20,21). These findings align with SCOT-HEART trial (n=1,430) demonstrating hsTnI improves pre-test probability estimation (OR 1.35, CI 1.25-1.46 per 2-fold increment) (22) and PROMISE (n=1,838) showing hsTnI independently predicts obstructive CAD presence and severity (OR 1.25 per IQR increase) (23), though neither specifically assessed incremental discriminative value beyond current ESC guideline-recommended diagnostic approaches or quantified category-specific benefits.

NT-proBNP emerged as the strongest mortality predictor across all patient categories, substantially outperforming hsTnT. This differential utility—hsTnT for diagnosis, NT-proBNP for prognosis—reflects distinct pathophysiology: hsTnT indicates injury relevant to obstruction detection, while NT-proBNP reflects ventricular stress, remodeling, and neurohumoral activation directly predicting mortality (3,4,24). NT-proBNP was the only biomarker maintaining prognostic discrimination across all therapeutic strategies. These findings contrast with the MICA study demonstrating hsTnI maintains independent prognostic value in stable angina patients (25). However, our results align with previous reports demonstrating robust MACE prediction by NT-proBNP in CCS patients with normal EF (26), while providing novel contributions: (1) treatment-interaction analysis demonstrating differential survival associations by NT-proBNP status and (2) 12.6-year median follow-up far exceeding typical 2–5-year investigations. Notably, biomarker-specific patterns emerged, with copeptin showing particularly strong performance in CABG patients. As a stable surrogate of arginine vasopressin release reflecting chronic neuroendocrine activation, copeptin’s superior discriminative capacity in this subgroup may indicate more advanced coronary disease burden with greater myocardial dysfunction (27).

The proposed 150 pg/mL NT-proBNP cutoff is supported by continuous spline analysis showing good discrimination at this level (Supplementary Figure S4). This post-hoc derived threshold is clinically meaningful, situated between ESC guideline thresholds for ruling out heart failure (125 pg/mL) and diagnosing acute heart failure (>300 pg/mL) (7), capturing early ventricular dysfunction and neurohumoral activation that precede overt heart failure (28). The significant interaction between NT-proBNP levels and revascularisation survival demonstrated differential prognostic patterns: patients with baseline NT-proBNP <150 pg/mL showed comparably low mortality irrespective of revascularisation. In contrast, NT-proBNP ≥150 pg/mL identified patients with substantially elevated baseline mortality (HR 5.75, p<0.001) and significant mortality reduction with revascularisation (HR 3.43, p<0.001), albeit with persistent residual risk. Importantly, revascularisation provides benefits beyond mortality including symptom relief and improved quality of life, which may be particularly relevant for patients with elevated NT-proBNP. Clinically, this threshold may inform risk stratification: patients with NT-proBNP ≥ 150 pg/mL appear to represent a higher-risk phenotype requiring intensified medical therapy, closer surveillance, and potentially novel therapeutic approaches (9,20,21). This biomarker-guided approach parallels NT-proBNP-guided heart failure therapy (7), though prospective validation is needed before clinical implementation.

Several limitations warrant consideration. The single-center observational design with single timepoint measurements precludes definitive causal inference and lacks external validation in independent cohorts, limiting generalizability. Most importantly, treatment allocation was clinician-directed. Despite rigorous PSM achieving good balance (mean |SMD| 0.069) with 19 confounders, unmeasured confounding cannot be excluded (Supplementary Table S4). Our analysis focused on mortality without assessing symptom relief, quality of life, or heart failure hospitalization—important revascularisation benefits particularly relevant for high-risk patients. We used invasive angiography with anatomical stenosis ≥50%—the established standard (29)—which demonstrates substantial discordance to functional relevance (30), potentially explaining moderate biomarker diagnostic performance. Fractional flow reserve and non-hyperemic pressure indices were not systematically performed. Exercise testing was available in 79.9% but did not add incremental diagnostic value over hsTnT (p=0.98) and showed no association with anatomic CAD presence (p=0.53). However, comprehensive sensitivity analyses using stricter anatomic thresholds (≥75%/LMCA≥50%, differing by only 5.4% from ≥50% cohort) demonstrated stable diagnostic and prognostic performance (Supplementary Table S5), supporting robustness of findings across stenosis severity definitions. Guideline-recommended non-invasive imaging now preferred after RF-CL assessment (2) was unavailable, precluding comparison with currently recommended modalities. Outcome assessment was limited by unavailable cause-specific mortality and missing ICD implantation data. Relatively small treatment subgroup samples (particularly CABG, n=267) may limit statistical power. The 150 pg/mL threshold was data-derived using optimization methods, risking overfitting. While internal bootstrap validation supports model stability, generalizability to other populations, assays, or time periods remains unproven. The threshold should be considered exploratory pending independent replication. Finally, while 12.6-year follow-up is substantial, contemporary therapeutic advances may alter the prognostic landscape.

In conclusion, this comprehensive biomarker analysis demonstrates differential diagnostic versus prognostic utility in CCS. High-sensitivity troponin T provides risk factor-weighted, clinical likelihood category-dependent diagnostic value following an inverse gradient pattern (greatest benefit in very low-risk patients), while NT-proBNP emerged as the universal prognostic biomarker. The 150 pg/mL NT-proBNP threshold identified two distinct phenotypes: patients below showed comparably low mortality regardless of revascularisation (HR 0.98, p=0.910), whereas patients above demonstrated elevated baseline mortality (HR 5.75, p<0.001) with 40% relative reduction following revascularisation (HR 3.43, p<0.001) but persistent 3.43-fold residual risk. Elevated NT-proBNP thus identifies a persistently high-risk phenotype requiring comprehensive management beyond anatomic intervention, including guideline-directed medical therapy, intensive risk factor modification, and close surveillance. Given the observational design with inherent confounding, these hypothesis-generating findings warrant prospective multicenter RCT validation to determine whether biomarker-guided strategies improve outcomes and cost-effectiveness in contemporary CCS management.

## Data Availability

The data that support the findings of this study are available from the corresponding author upon reasonable request. Due to privacy and ethical restrictions, individual participant data cannot be made publicly available. Requests for data access should be directed to the Leipzig Heart Study steering committee and are subject to approval by the local ethics committee and compliance with data protection regulations.

## ACKNOWLEDGEMENTS

The authors acknowledge all participants of the LIFE-Heart study for spending their time and donating blood, as well as for their continued participation in our follow-up endeavours. We thank Annegret Unger and Kay Olischer for running the study ambulance, and Ronny Dathe for database development.

## AUTHORS’ CONTRIBUTIONS

AT and JN conceived and designed the study. AT, JT and HT contributed to data acquisition. AT, HK, and MS performed statistical analysis. AT drafted the manuscript. All authors (AT, JN, JT, HT, CS, DL, TL, AD, SH, HK, MS, TK) contributed to data interpretation, critically revised the manuscript for important intellectual content, and approved the final version for submission. AT takes responsibility for the integrity of the data and the accuracy of the data analysis.

## NON-STANDARD ABBREVIATIONS AND ACRONYMS

ATC: Anatomical Therapeutic Chemical (classification system)
AUC: Area Under the Curve
CABG: Coronary Artery Bypass Grafting
CAD: Coronary Artery Disease
CCS: Chronic Coronary Syndrome
CKD-EPI: Chronic Kidney Disease Epidemiology Collaboration
CRF: Cardiovascular Risk Factor (model)
DCA: Decision Curve Analysis
eGFR: Estimated Glomerular Filtration Rate
ESC: European Society of Cardiology
HDL-C: High-density Lipoprotein Cholesterol
HR: Hazard Ratio
hsCRP: High-sensitivity C-reactive Protein
hsTnT: High-sensitivity Cardiac Troponin T
IL-6: Interleukin-6
IQR: Interquartile Range
LDL-C: Low-density Lipoprotein Cholesterol
LIFE-Heart: Leipzig Research Centre for Civilisation Diseases – Heart Study
NT-proBNP: N-terminal pro-B-type Natriuretic Peptide
NYHA: New York Heart Association
OMT: Optimal Medical Therapy
PCI: Percutaneous Coronary Intervention
PSM: Propensity Score Matching
PTP: Pre-test Probability
RF-CL: Risk Factor-weighted Clinical Likelihood
ROC: Receiver Operating Characteristic
SMD: Standardized Mean Difference
TG: Triglycerides
WHR: Waist-hip Ratio

---

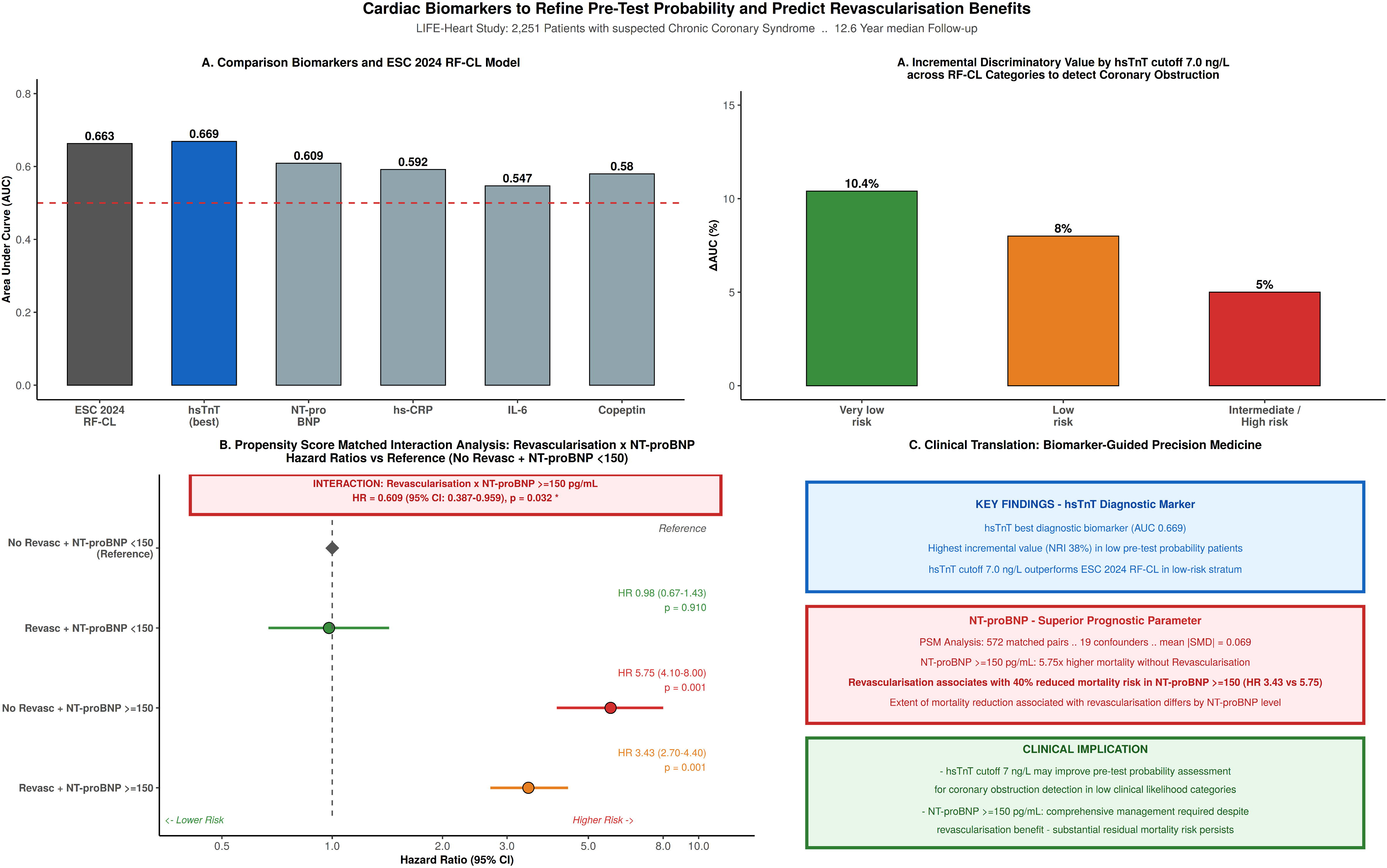

## REFERENCES

1. Knuuti J, Wijns W, Saraste A, Capodanno D, Barbato E, Funck-Brentano C, Prescott E, Storey RF, Deaton C, Cuisset T, et al. 2019 ESC Guidelines for the diagnosis and management of chronic coronary syndromes. Eur Heart J. 2020;41:407–477.

2. Vrints C, Andreotti F, Koskinas KC, Rossello X, Adamo M, Ainslie J, Banning AP, Budaj A, Buechel RR, Chiariello GA, et al. 2024 ESC Guidelines for the management of chronic coronary syndromes. Eur Heart J. 2024;45:3415–3537.

3. Biomarkers Definitions Working Group. Biomarkers and surrogate endpoints: preferred definitions and conceptual framework. Clin Pharmacol Ther. 2001;69:89–95.

4. Morrow DA, de Lemos JA. Benchmarks for the assessment of novel cardiovascular biomarkers. Circulation. 2007;115:949–952.

5. Thygesen K, Alpert JS, Jaffe AS, Chaitman BR, Bax JJ, Morrow DA, White HD; ESC Scientific Document Group. Fourth universal definition of myocardial infarction (2018). Eur Heart J. 2019;40:237–269.

6. Chacko L, P Howard J, Rajkumar C, Nowbar AN, Kane C, Mahdi D, Foley M, Shun-Shin M, Cole G, Sen S, et al. Effects of percutaneous coronary intervention on death and myocardial infarction stratified by stable and unstable coronary artery disease: A meta-analysis of randomized controlled trials. Circ Cardiovasc Qual Outcomes. 2020;13:e006363.

7. McDonagh TA, Metra M, Adamo M, Gardner RS, Baumbach A, Böhm M, Burri H, Butler J, Čelutkienė J, Chioncel O, et al. 2021 ESC Guidelines for the diagnosis and treatment of acute and chronic heart failure. Eur Heart J. 2021;42:3599–3726.

8. Pencina MJ, D’Agostino RB, Steyerberg EW. Extensions of net reclassification improvement calculations to measure usefulness of new biomarkers. Stat Med. 2011;30:11–21.

9. Ridker PM, Everett BM, Thuren T, MacFadyen JG, Chang WH, Ballantyne C, Fonseca F, Nicolau J, Koenig W, Anker SD, et al. Antiinflammatory therapy with canakinumab for atherosclerotic disease. N Engl J Med. 2017;377:1119–1131.

10. Maron DJ, Hochman JS, Reynolds HR, Bangalore S, O’Brien SM, Boden WE, Chaitman BR, Senior R, López-Sendón J, Alexander KP, et al. Initial invasive or conservative strategy for stable coronary disease. N Engl J Med. 2020;382:1395–1407.

11. Panuccio G, Carabetta N, Torella D, De Rosa S. Percutaneous coronary revascularization versus medical therapy in chronic coronary syndromes: An updated meta-analysis of randomized controlled trials. Eur J Clin Invest. 2024;54:e14303.

12. Scholz M, Henger S, Beutner F, Teren A, Baber R, Willenberg A, Ceglarek U, Pott J, Burkhardt R, Thiery J. Cohort Profile: The Leipzig Research Center for Civilization Diseases-Heart Study (LIFE-Heart). Int J Epidemiol. 2020;49:1439–1440h.

13. Gensini GG. A more meaningful scoring system for determining the severity of coronary heart disease. Am J Cardiol. 1983;51:606.

14. Morgenthaler NG, Struck J, Alonso C, Bergmann A. Assay for the measurement of Copeptin, a stable C-terminal vasopressin fragment in human plasma. Clin Chem. 2006;52:112–119.

15. DeLong ER, DeLong DM, Clarke-Pearson DL. Comparing the areas under two or more correlated receiver operating characteristic curves: a nonparametric approach. Biometrics. 1988;44:837–845.

16. Youden WJ. Index for rating diagnostic tests. Cancer. 1950;3:32–35.

17. Netto J, Teren A, Burkhardt R, Willenberg A, Beutner F, Henger S, Schuler G, Thiele H, Isermann B, Thiery J, et al. Biomarkers for non-invasive stratification of coronary artery disease and prognostic impact on long-term survival in patients with stable coronary heart disease. Nutrients. 2022;14:3433.

18. Apple FS, Collinson PO; IFCC Task Force on Clinical Applications of Cardiac Biomarkers. Analytical characteristics of high-sensitivity cardiac troponin assays. Clin Chem. 2012;58:54–61.

19. Omland T, de Lemos JA, Sabatine MS, Christophi CA, Rice MM, Jablonski KA, Tjora S, Domanski MJ, Gersh BJ, Rouleau JL, et al. A sensitive cardiac troponin T assay in stable coronary artery disease. N Engl J Med. 2009;361:2538–2547.

20. Tardif JC, Kouz S, Waters DD, Bertrand OF, Diaz R, Maggioni AP, Pinto FJ, Ibrahim R, Gamra H, Kiwan GS, Berry C, et al. Efficacy and safety of low-dose colchicine after myocardial infarction. N Engl J Med. 2019;381:2497–2505.

21. Leopold JA, Loscalzo J. Emerging role of precision medicine in cardiovascular disease. Circ Res. 2018;122:1302–1315.

22. Adamson PD, Hunter A, Madsen DM, Shah ASV, McAllister DA, Pawade TA, Williams MC, Berry C, Boon NA, Flather M, et al. High-Sensitivity Cardiac Troponin I and the Diagnosis of Coronary Artery Disease in Patients With Suspected Angina Pectoris. Circ Cardiovasc Qual Outcomes. 2018;11:e004227.

23. Januzzi JL Jr, Suchindran S, Coles A, Ferencik M, Patel MR, Hoffmann U, Ginsburg GS, Douglas PS; PROMISE Investigators. High-Sensitivity Troponin I and Coronary Computed Tomography in Symptomatic Outpatients With Suspected CAD: Insights From the PROMISE Trial. JACC Cardiovasc Imaging. 2019 Jun;12:1047–1055

24. Zethelius B, Berglund L, Sundström J, Ingelsson E, Basu S, Larsson A, Venge P, Arnlöv J. Use of multiple biomarkers to improve the prediction of death from cardiovascular causes. N Engl J Med. 2008 May 15;358:2107–16.

25. Wereski R, Adamson P, Shek Daud NS, McDermott M, Taggart C, Bularga A, Kimenai DM, Lowry MTH, Tuck C, Anand A, et al. High-Sensitivity Cardiac Troponin for Risk Assessment in Patients With Chronic Coronary Artery Disease. J Am Coll Cardiol. 2023;82:473–485.

26. Liu HH, Cao YX, Jin JL, Guo YL, Zhu CG, Wu NQ, Gao Y, Zhang Y, Xu RX, Dong Q, et al. Prognostic value of NT-proBNP in patients with chronic coronary syndrome and normal left ventricular systolic function according to glucose status: a prospective cohort study. Cardiovasc Diabetol. 2021;20:84.

27. Deveci OS, Ozmen C, Karaaslan MB, Celik AI. Could Serum Copeptin Level Be an Indicator of Coronary Artery Disease Severity in Patients with Unstable Angina? Int Heart J. 2021 May 29;62528–533.

28. Bibbins-Domingo K, Gupta R, Na B, Wu AH, Schiller NB, Whooley MA. N-terminal fragment of the prohormone brain-type natriuretic peptide (NT-proBNP), cardiovascular events, and mortality in patients with stable coronary heart disease. JAMA. 2007 Jan 10;297:169–76.

29. Rosenthal RL. The 50% coronary stenosis. Am J Cardiol. 2015;115:1162–1165.

30. Toth G, Hamilos M, Pyxaras S, Mangiacapra F, Nelis O, De Vroey F, Di Serafino L, Muller O, Van Mieghem C, Wyffels E, et al. Evolving concepts of angiogram: fractional flow reserve discordances in 4000 coronary stenoses. Eur Heart J. 2014;35:2831–2838.

