## Supplemental Material for "Cardiac Biomarkers to Refine Pre-Test Probability for Coronary Obstruction and Predict Survival after Revascularisation in Chronic Coronary Syndrome"

**Supplementary Figure S1**

**Assessment of Proportional Hazards Assumption for Cox Regression Models**

Schoenfeld residual plots (left panels) and log-log survival curves (right panels) for all biomarker Cox regression models. Each row represents one biomarker: N-terminal pro-B-type natriuretic peptide (NT-proBNP), high-sensitivity troponin T (hsTnT), high-sensitivity C-reactive protein (hsCRP), interleukin-6 (IL-6), and copeptin.

Left panels show standardized Schoenfeld residuals plotted against follow-up time, with red LOWESS smoothed trend lines and gray reference lines at zero. Spearman correlation (ρ) between residuals and time tests for monotonic trends; non-significant correlations (p >0.05, indicated by ✓ and green background) support the proportional hazards assumption. Right panels display log(-log(survival)) versus log(time) for biomarker tertiles; parallel curves indicate proportional hazards.

Results: hsTnT (ρ = -0.063, p = 0.101) and hsCRP (ρ = 0.062, p = 0.107) satisfied proportional hazards. NT-proBNP (ρ = -0.211, p <0.001), IL-6 (ρ = 0.265, p <0.001), and copeptin (ρ = -0.129, p = 0.001) showed statistically significant but modest deviations (all |ρ| <0.3), consistent with biologically plausible time-varying effects over 11.6-year mean follow-up. Log-log curves show generally parallel trends for all biomarkers. The primary 4-group interaction analysis remains valid as it does not assume constant hazard ratios across NT-proBNP strata.

*Abbreviations: hsCRP, high-sensitivity C-reactive protein; hsTnT, high-sensitivity troponin T; IL-6, interleukin-6; LOWESS, locally weighted scatterplot smoothing; NT-proBNP, N-terminal pro-B-type natriuretic peptide.*

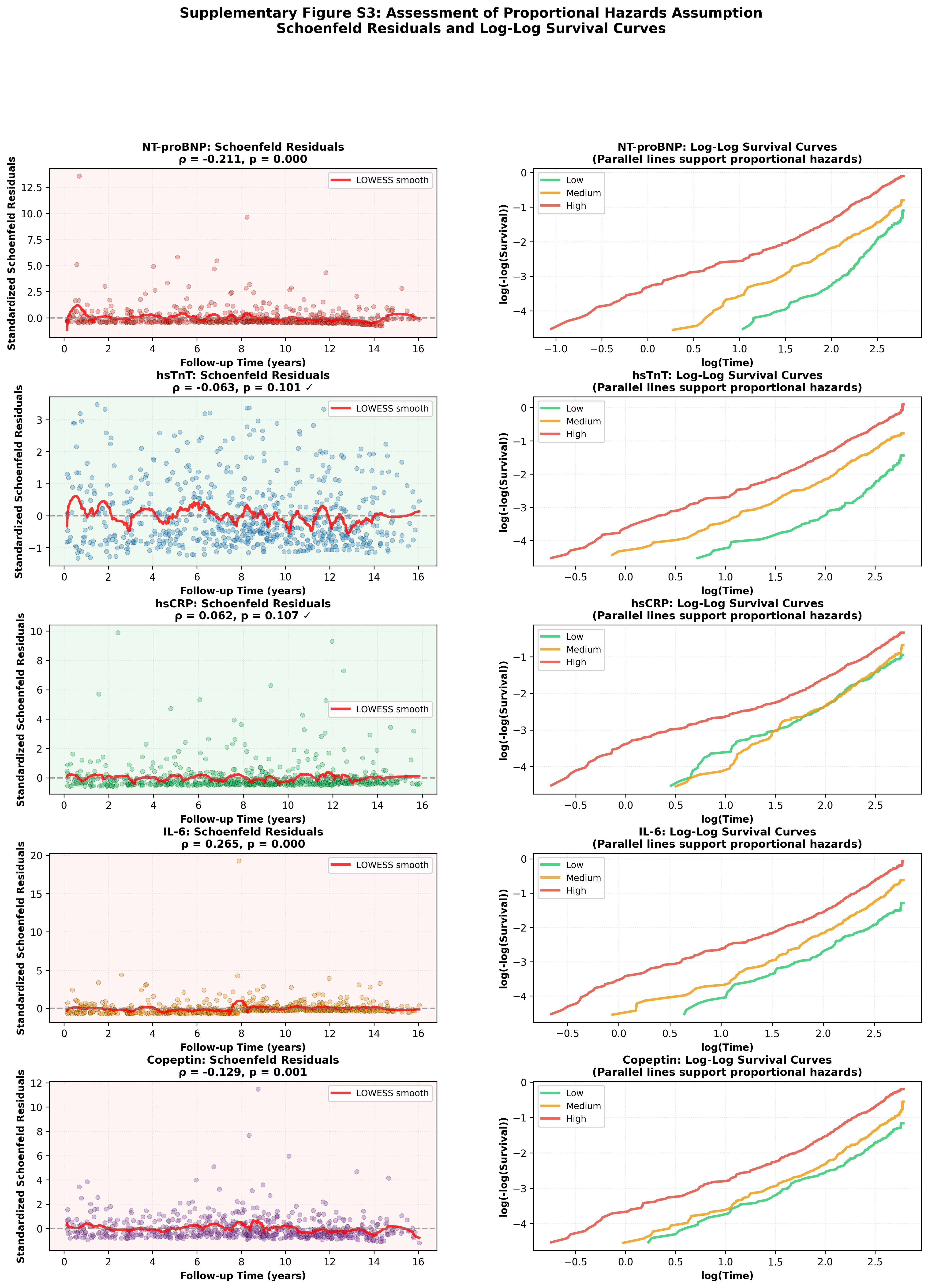

**Supplementary Figure S2**

**Propensity Score Matching Quality Assessment – Love Plot**

SMD for 19 confounders before (red circles) and after (blue circles) 1:1 nearest neighbor PSM without replacement (caliper 0.2 SD). Analysis restricted to patients with obstructive CAD and NT-proBNP ≥150 pg/mL (N=1,144 matched, 572 pairs). The 19 confounders include: age, sex, BMI, waist-hip ratio, smoking status, diabetes mellitus, history of hyperlipidemia, history of premature CAD, statin use, antihypertensive medication, eGFR, LDL cholesterol, HDL cholesterol, triglycerides, systolic BP, diastolic BP, left-ventricular ejection fraction, Gensini score, and number of major epicardial coronary vessels with ≥ one stenosis ≥50%. Green shaded zone indicates excellent balance (|SMD|<0.1). Connecting gray lines show balance improvement. Pre-matching: mean |SMD|=0.226. Post-matching: mean |SMD|=0.069, representing 69.5% reduction. All confounders achieve |SMD|<0.15; 15/19 (78.9%) achieve Austin criterion (|SMD|<0.1). Critical anatomic confounders excellently balanced: Gensini score |SMD|=0.028, number of stenoses |SMD|=0.043.

*Abbreviations: BMI, body mass index; BP, blood pressure; CAD, coronary artery disease; eGFR, estimated glomerular filtration rate; HDL, high-density lipoprotein; LDL, low-density lipoprotein; LVEF, left ventricular ejection fraction; NT-proBNP, N-terminal pro-B-type natriuretic peptide; PSM, propensity score matching; SD, standard deviation; SMD, standardized mean difference.*

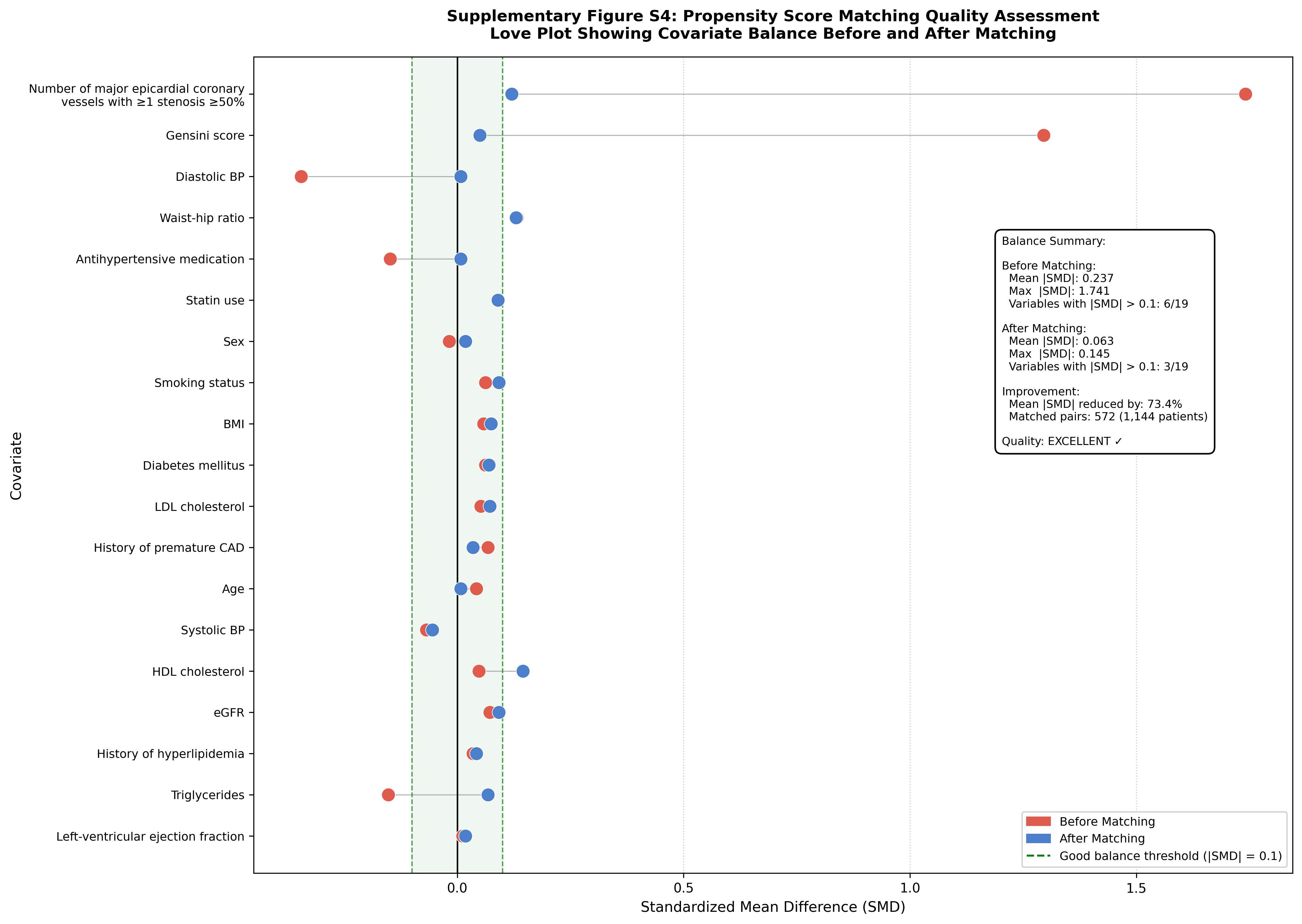

### Supplementary Figure S3.

**Calibration and Clinical Utility Analysis for Obstructive CAD Detection.**

**(A)** Calibration curves showing agreement between predicted probabilities and observed proportions of obstructive CAD for ESC 2024 RF-CL model alone (blue) versus combined model with hsTnT (red). The RF-CL model demonstrated acceptable calibration (Hosmer-Lemeshow χ²=12.4, p=0.134; Brier score=0.224), with addition of hsTnT ≥7.0 ng/L cutoff improving calibration (Hosmer-Lemeshow χ²=8.9, p=0.351; Brier score=0.216; ΔBrier=-0.008, p<0.001).

**(B)** Decision Curve Analysis comparing net benefit of ESC 2024 RF-CL alone (blue) versus combined with hsTnT (red) across threshold probabilities for obstructive CAD. The combined model demonstrates superior net benefit across 20-60% threshold probability range, with maximum incremental benefit at 30-45% thresholds. Gray lines represent "treat all" and "treat none" strategies.

*Abbreviations: CAD, coronary artery disease, ESC, European Society of Cardiology; RF-CL, risk factor weighted clinical likelihood; hsTnT, high-sensitivity Troponin T.*

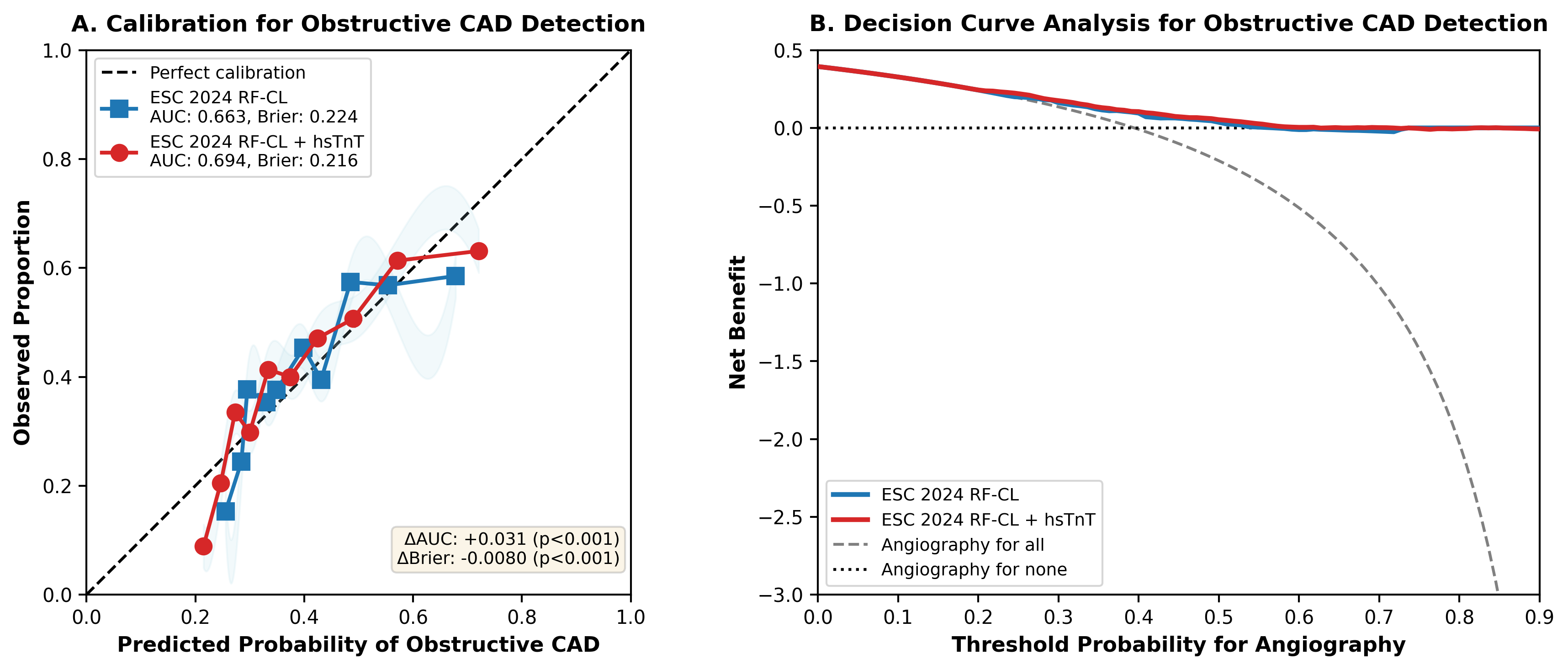

**Supplementary Figure S4**

**Continuous NT-proBNP × Revascularization Interaction – Restricted Cubic Spline Analysis**

Continuous modeling of NT-proBNP × revascularization interaction using restricted cubic splines with 4 knots placed at 5th, 35th, 65th, and 95th percentiles of log-transformed NT-proBNP. Hazard ratios for revascularization (versus optimal medical therapy) are shown across the NT-proBNP spectrum in patients with obstructive coronary artery disease (≥50% stenosis). Blue line represents point estimates with 95% confidence intervals (shaded area). Red circles indicate empirical hazard ratios at specific NT-proBNP levels. Red dashed vertical line marks the 150 pg/mL threshold used in primary dichotomous analyses.

Models adjusted for cardiovascular risk factors including Gensini score. Overall spline interaction test: likelihood ratio statistic = 7.516 (df=3), p = 0.0571 (borderline significance). Analysis included 1,921 patients with 513 deaths. The continuous model demonstrates a dose-response relationship with strongest revascularization benefit at lowest NT-proBNP levels (HR 0.50 at 50 pg/mL), crossing null effect around 150 pg/mL, and diminishing benefit at higher levels. This pattern supports biological plausibility of NT-proBNP effect modification despite conservative continuous modeling approach.

*Abbreviations: CAD, coronary artery disease; df, degrees of freedom; HR, hazard ratio; LR, likelihood ratio; NT-proBNP, N-terminal pro-B-type natriuretic peptide; OMT, optimal medical therapy.*

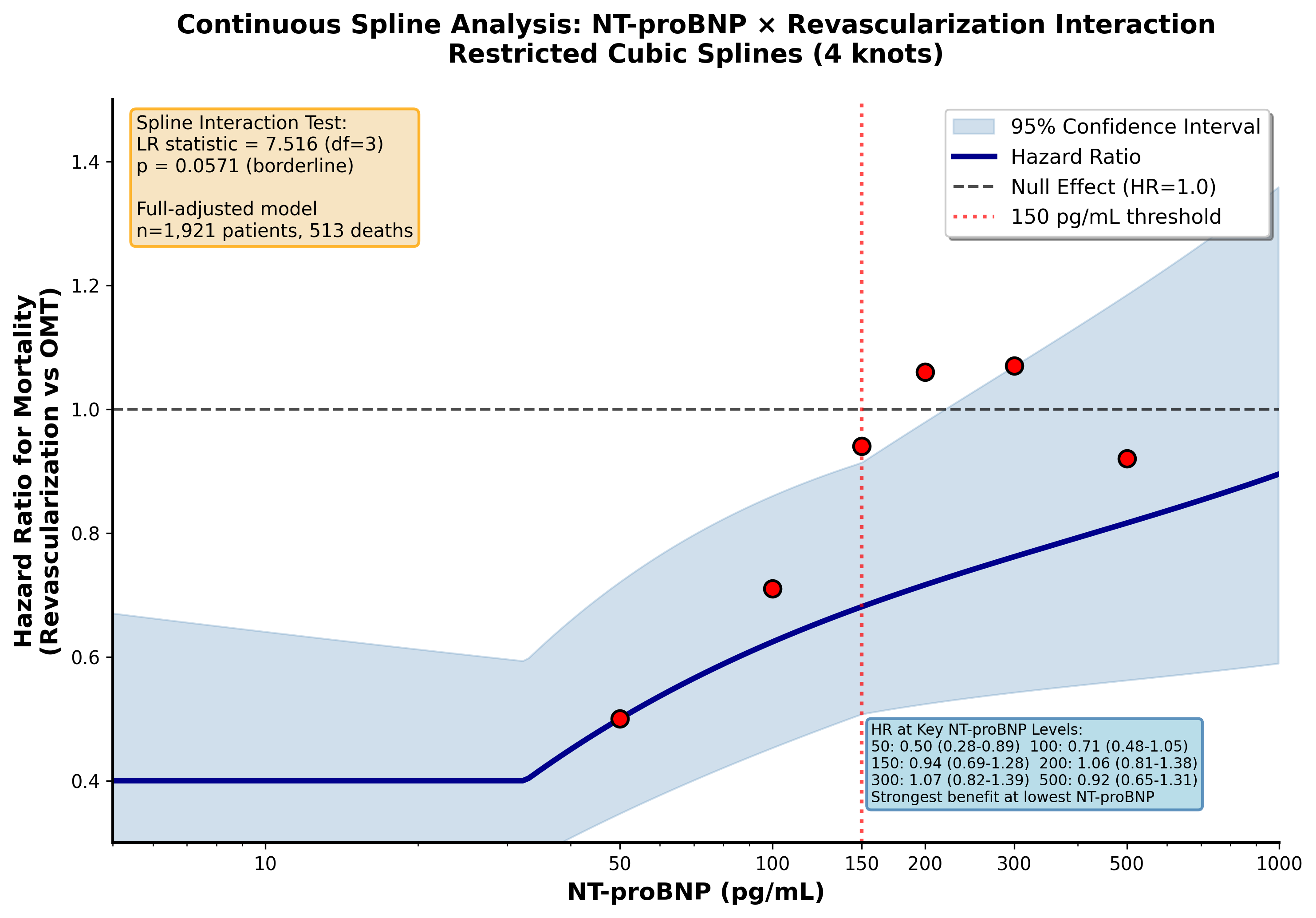

### Supplementary Table S1.

**Discrimination Improvement with hsTnT ≥7.0 ng/L Cutoff by ESC 2024 RF-CL Category.** Incremental diagnostic value stratified by baseline pre-test probability categories. Bootstrap validation with 500 iterations.

*Abbreviations: CAD, coronary artery disease, ESC, European Society of Cardiology; RF-CL, risk factor weighted clinical likelihood; hsTnT, high-sensitivity Troponin T; AUC, area under the curve.*

| Metric | Value | 95% CI | p-value |
| --- | --- | --- | --- |
| Sample & Outcome |  |  |  |
| Total sample size | 2,251 | – | – |
| Obstructive CAD | 888 (39.4%) | – | – |
| Optimal Cutoff |  |  |  |
| Optimal hsTnT cutoff | 7.0 ng/L | – | – |
| Youden Index | 0.264 | 0.223-0.299 | – |
| Sensitivity | 72.4% | 69.4-75.4% | – |
| Specificity | 54.0% | 51.3-56.5% | – |
| Discrimination (Obstructive CAD) |  |  |  |
| AUC (ESC 2024 RF-CL alone) | 0.663 | 0.639-0.686 | – |
| AUC (ESC 2024 RF-CL + hsTnT) | 0.694 | 0.671-0.717 | – |
| ΔAUC | +0.031 | -0.001 to +0.064 | <0.001 |
| Calibration (Obstructive CAD) |  |  |  |
| Brier Score (ESC 2024 RF-CL) | 0.224 | 0.219-0.229 | – |
| Brier Score (ESC 2024 RF-CL + hsTnT) | 0.216 | 0.211-0.221 | – |
| ΔBrier Score | -0.008 | -0.013 to -0.003 | <0.001 |

### Supplementary Table S2.

**Detailed reclassification matrix by hsTnT 7.0 ng/L cutoff to detect obstructive CAD across RF-CL categories.** Complete documentation of patient reclassification patterns showing movement between risk categories. Rows represent baseline RF-CL risk classification; columns represent reclassification after adding hsTnT ≥7.0 ng/L cutoff to the model.

*Abbreviations: CAD, coronary artery disease, RF-CL, risk factor weighted clinical likelihood; hsTnT, high-sensitivity Troponin T; AUC, area under the curve.*

**Reclassification Logic:**

- If hsTnT ≥7.0 ng/L: risk increases by 1 category (Category 1→2, Category 2→3, Category 3→3)
- If hsTnT <7.0 ng/L: risk decreases by 1 category (Category 1→1, Category 2→1, Category 3→2)
- Note: Category 2 patients always move to either Category 1 or 3 (none remain in Category 2)

**A. Patients with Obstructive CAD (n=888):**

| Baseline RF-CL | Reclassified: Very Low (1) | Reclassified: Low (2) | Reclassified: Intermediate/High (3) | Total |
| --- | --- | --- | --- | --- |
| Very Low (1) | 21 (44.7%) | 26 (55.3%) | 0 (0.0%) | 47 |
| Low (2) | 121 (38.7%) | 0 (0.0%) | 192 (61.3%) | 313 |
| Intermediate/High (3) | 0 (0.0%) | 107 (20.3%) | 421 (79.7%) | 528 |
| Total Reclassified | 142 | 133 | 613 | 888 |

**Movement Summary:**

- Upward reclassification: 218 patients (24.5%) - moved to higher risk category
- Downward reclassification: 228 patients (25.7%) - moved to lower risk category
- No change: 442 patients (49.8%) - remained in same category
- False negatives (obstructive CAD with hsTnT <7.0 ng/L): 249 patients (28.0%)

**B. Patients without Obstructive CAD (n=1,363):**

| Baseline RF-CL | Reclassified: Very Low (1) | Reclassified: Low (2) | Reclassified: Intermediate/High (3) | Total |
| --- | --- | --- | --- | --- |
| Very Low (1) | 216 (83.1%) | 44 (16.9%) | 0 (0.0%) | 260 |
| Low (2) | 369 (57.9%) | 0 (0.0%) | 268 (42.1%) | 637 |
| Intermediate/High (3) | 0 (0.0%) | 152 (32.6%) | 314 (67.4%) | 466 |
| Total Reclassified | 585 | 196 | 582 | 1,363 |

**Movement Summary:**

- Upward reclassification: 312 patients (22.9%) - moved to higher risk category
- Downward reclassification: 521 patients (38.2%) - moved to lower risk category
- No change: 530 patients (38.9%) - remained in same category
- True negatives benefit: More downward (38.2%) than upward (22.9%) movement demonstrates appropriate risk reduction in patients without obstructive CAD

**Clinical Interpretation:**

The reclassification patterns demonstrate that hsTnT ≥7.0 ng/L cutoff provides meaningful risk stratification: (1) In patients WITH obstructive CAD, nearly balanced upward (24.5%) and downward (25.7%) movements indicate that while hsTnT adds diagnostic value, it is not perfectly discriminative; (2) In patients WITHOUT obstructive CAD, substantially more downward (38.2%) than upward (22.9%) reclassification demonstrates that low hsTnT appropriately identifies lower-risk patients; (3)

### Supplementary Table S3

**Safety Analysis: Survival of False Negative Patients.** Comparison of mortality over 12.6-year follow-up in patients with obstructive CAD but hsTnT <7.0 ng/L ("false negatives") versus other patients within their respective reclassified risk categories. False negatives include patients who remained in or were reclassified to lower risk categories despite having obstructive CAD. Unadjusted hazard ratios from Cox regression with false negative status as sole predictor; RF-CL adjusted hazard ratios from Cox regression adjusted for RF-CL variables.

*Abbreviations: CAD, coronary artery disease; hsTnT, high-sensitivity Troponin T; RF-CL, risk factor weighted clinical likelihood.*

| Reclassified Category | False Negatives (n) | Reference Group (n) | Deaths in False Negatives (%) | Unadjusted HR (95% CI) | RF-CL Adjusted HR (95% CI) |
| --- | --- | --- | --- | --- | --- |
| Very Low | 142 | 585 | 26 (18.3%) | 1.83 (1.16-2.89) p=0.009 | 1.29 (0.81-2.06) p=0.283 |
| Low | 107 | 222 | 22 (20.6%) | 0.98 (0.59-1.63) p=0.953 | 0.90 (0.53-1.54) p=0.707 |
| Overall | 249 | 807 | 48 (19.3%) | 1.52 (1.08-2.13) p=0.016 | 1.10 (0.77-1.56) p=0.608 |

False Negatives = patients with obstructive CAD but hsTnT <7.0 ng/L

Reference Group = All other patients in the same reclassified risk category

RF-CL variables: age, sex, diabetes mellitus, current smoking, hypertension, dyslipidemia, family history of premature CAD

**Detailed Breakdown of False Negatives:**

- Very Low category (n=142): Includes 21 patients who stayed in Category 1 (baseline Cat 1 → Cat 1) and 121 patients who moved down from Category 2 to Category 1 (baseline Cat 2 → Cat 1)
- Low category (n=107): Patients who moved down from Category 3 to Category 2 (baseline Cat 3 → Cat 2)
- Overall (n=249): All patients with obstructive CAD and hsTnT <7.0 ng/L in reclassified Categories 1 and 2

**Clinical Interpretation:**

Unadjusted analysis revealed that false negative patients had significantly higher mortality in the Very Low category (HR=1.83, p=0.009) and overall (HR=1.52, p=0.016). However, after adjustment for RF-CL variables, this apparent excess mortality was substantially attenuated and no longer statistically significant across all categories: Very Low (HR=1.29, p=0.283), Low (HR=0.90, p=0.707), and Overall (HR=1.10, p=0.608). RF-CL factors explained 65.0% of excess risk in the Very Low category and 81.3% overall. This pattern demonstrates that the apparent unadjusted mortality differences were primarily attributable to baseline cardiovascular risk factor burden rather than low hsTnT itself.

### Supplementary Table S4

**Cardiovascular Risk Factor Distribution by Treatment Group and NT-proBNP Status**

Baseline characteristics stratified by treatment category (no/non-obstructive CAD, optimal medical therapy, percutaneous coronary intervention, coronary artery bypass grafting) and further dichotomized by NT-proBNP threshold of 150 pg/mL. Within each treatment group, p-values compare patients with NT-proBNP <150 pg/mL versus ≥150 pg/mL using Mann-Whitney U test for continuous variables and chi-square test for categorical variables. Continuous variables presented as median (IQR); categorical variables as N (%).

*Abbreviations: CAD, coronary artery disease; oCCS, obstructive chronic coronary syndrome;IQR, inter-quartal range; OMT, optimal medical therapy; PCI, percutaneous coronary intervention; CABG, coronary artery bypass grafting; NT-proBNP, N-terminal pro-B-type natriuretic peptide; BMI, body mass index; MI, myocardial infarction; WHR, waist-hip ratio; LDL, low-density lipoprotein; HDL, high-density lipoprotein; eGFR, estimated glomerular filtration rate.*

| Characteristics | No/Non-oCCS BNP<150 N/Median | %/IQR | No/Non-oCCS BNP≥150 N/Median | %/IQR | p-value | OMT BNP<150 N/Median | %/IQR | OMT BNP≥150 N/Median | %/IQR | p-value |
| --- | --- | --- | --- | --- | --- | --- | --- | --- | --- | --- |
| N | 823 |  | 540 |  | - | 115 |  | 162 |  | - |
| Female sex (N) | 336 | 40.8% | 269 | 49.8% | 0.001 | 20 | 17.4% | 36 | 22.2% | 0.404 |
| Age (years) | 58.38 | 51.56-66.56 | 67.44 | 59.45-72.90 | <.001 | 61.97 | 55.16-69.69 | 67.48 | 58.80-72.49 | <.001 |
| BMI (kg/m²) | 29.27 | 26.28-32.62 | 29.41 | 26.63-33.21 | 0.333 | 29.29 | 26.12-33.14 | 29.27 | 26.03-32.81 | 0.832 |
| Waist-hip ratio | 0.97 | 0.89-1.03 | 0.94 | 0.87-1.02 | 0.001 | 1.02 | 0.97-1.06 | 1.01 | 0.94-1.05 | 0.175 |
| Diabetes mellitus (N) | 187 | 22.7% | 148 | 27.4% | 0.057 | 39 | 33.9% | 58 | 35.8% | 0.844 |
| Family history with MI (N) | 117 | 14.2% | 58 | 10.7% | 0.073 | 18 | 15.7% | 18 | 11.1% | 0.354 |
| Current smoker (N) | 455 | 55.3% | 262 | 48.5% | 0.017 | 85 | 73.9% | 114 | 70.4% | 0.610 |
| Antihypertensive medication (N) | 644 | 78.3% | 500 | 92.6% | <.001 | 99 | 86.1% | 145 | 89.5% | 0.498 |
| Lipid lowering medication (N) | 263 | 32.0% | 196 | 36.3% | 0.110 | 57 | 49.6% | 79 | 48.8% | 0.993 |
| LDL-cholesterol (mmol/l) | 3.34 | 2.77-3.96 | 3.14 | 2.43-3.84 | <.001 | 3.32 | 2.71-4.17 | 3.17 | 2.50-4.06 | 0.183 |
| HDL-cholesterol (mmol/l) | 1.32 | 1.11-1.60 | 1.33 | 1.07-1.65 | 0.848 | 1.25 | 1.00-1.52 | 1.24 | 0.98-1.51 | 0.985 |
| Triglycerides (mmol/l) | 1.68 | 1.15-2.50 | 1.51 | 1.09-2.21 | 0.007 | 2.01 | 1.42-2.76 | 1.67 | 1.27-2.33 | 0.013 |
| eGFR (ml/min/1.73 m²) | 91.09 | 79.73-99.39 | 82.30 | 67.60-92.65 | <.001 | 90.05 | 78.22-99.88 | 84.11 | 69.02-93.53 | 0.003 |
| Ejection fraction (%) | 62.00 | 58.00-66.00 | 59.00 | 50.00-65.00 | <.001 | 62.00 | 57.00-66.00 | 57.00 | 48.00-64.00 | <.001 |
| N of coronary vessels with ≥50% stenosis | 0.0 | 0.0-1.0 | 0.0 | 0.0-1.0 | 0.002 | 2.0 | 2.0-3.0 | 2.0 | 2.0-3.0 | 0.539 |
| Gensini Score | 0.00 | 0.00-1.00 | 0.00 | 0.00-1.50 | 0.003 | 12.00 | 4.12-33.00 | 20.00 | 6.50-36.62 | 0.110 |

| Characteristics | PCI BNP<150 N/Median | %/IQR | PCI BNP≥150 N/Median | %/IQR | p-value | CABG BNP<150 N/Median | %/IQR | CABG BNP≥150 N/Median | %/IQR | p-value |
| --- | --- | --- | --- | --- | --- | --- | --- | --- | --- | --- |
| N | 184 |  | 160 |  | - | 100 |  | 167 |  | - |
| Female sex (N) | 42 | 22.8% | 58 | 36.2% | 0.009 | 9 | 9.0% | 28 | 16.8% | 0.111 |
| Age (years) | 61.28 | 55.67-69.77 | 68.16 | 60.55-72.62 | <.001 | 60.93 | 53.06-67.42 | 67.94 | 58.86-72.03 | <.001 |
| BMI (kg/m²) | 29.00 | 26.85-32.30 | 30.08 | 26.87-32.67 | 0.362 | 28.54 | 26.92-31.38 | 29.06 | 25.86-32.39 | 0.938 |
| Waist-hip ratio | 1.00 | 0.95-1.04 | 0.99 | 0.92-1.05 | 0.485 | 1.02 | 0.98-1.05 | 1.02 | 0.95-1.05 | 0.371 |
| Diabetes (N) | 52 | 28.3% | 67 | 41.9% | 0.011 | 27 | 27.0% | 59 | 35.3% | 0.203 |
| Family history with MI (N) | 29 | 15.8% | 23 | 14.4% | 0.836 | 23 | 23.0% | 22 | 13.2% | 0.057 |
| Current smoker (N) | 107 | 58.2% | 98 | 61.3% | 0.636 | 65 | 65.0% | 119 | 71.3% | 0.351 |
| Antihypertensive medication (N) | 158 | 85.9% | 146 | 91.2% | 0.166 | 84 | 84.0% | 144 | 86.2% | 0.749 |
| Lipid lowering medication (N) | 73 | 39.7% | 81 | 50.6% | 0.054 | 42 | 42.0% | 85 | 50.9% | 0.200 |
| LDL-cholesterol (mmol/l) | 3.71 | 2.89-4.51 | 3.23 | 2.31-3.93 | <.001 | 3.50 | 2.89-4.45 | 3.44 | 2.59-4.28 | 0.184 |
| HDL-cholesterol (mmol/l) | 1.22 | 1.01-1.38 | 1.23 | 1.02-1.50 | 0.256 | 1.21 | 1.03-1.37 | 1.20 | 0.99-1.52 | 0.682 |
| Triglycerides (mmol/l) | 1.95 | 1.39-2.87 | 1.70 | 1.25-2.25 | 0.002 | 1.83 | 1.35-2.48 | 1.66 | 1.25-2.37 | 0.300 |
| eGFR (ml/min/1.73 m²) | 85.94 | 75.62-95.95 | 79.59 | 66.29-91.41 | <.001 | 87.84 | 78.50-96.77 | 81.32 | 66.91-92.04 | 0.002 |
| Ejection fraction (%) | 63.00 | 59.00-66.00 | 59.00 | 52.00-65.00 | <.001 | 61.00 | 57.00-67.00 | 57.00 | 45.00-64.00 | <.001 |
| N of coronary vessels with ≥50% stenosis | 3.0 | 2.0-3.0 | 3.0 | 2.0-3.0 | 0.055 | 4.0 | 3.0-4.0 | 4.0 | 3.0-4.0 | 0.133 |
| Gensini Score | 20.00 | 12.00-32.62 | 26.00 | 14.00-42.88 | 0.019 | 48.00 | 31.75-70.50 | 59.00 | 42.00-81.00 | 0.003 |

**Key Findings:**

- Patient distribution: No/Non-obstructive CAD (n=1,363: BNP<150 n=823, BNP≥150 n=540), OMT (n=277: BNP<150 n=115, BNP≥150 n=162), PCI (n=344: BNP<150 n=184, BNP≥150 n=160), CABG (n=267: BNP<150 n=100, BNP≥150 n=167)
- Elevated NT-proBNP was consistently associated with older age (all p<0.001), lower ejection fraction (all p<0.001), and reduced renal function (all p≤0.003) across all treatment groups
- Traditional cardiovascular risk factors (diabetes, smoking status, lipid-lowering medication, LDL-cholesterol) showed relatively balanced distribution across NT-proBNP strata (most p>0.05)
- Gensini score was higher in elevated NT-proBNP subgroups (p<0.02 for all CAD subgroup except for obstructive CAD patients treated with OMT alone), indicating greater anatomical CAD complexity associated with elevated NT-proBNP values

### Supplementary Table S5

**Sensitivity Analysis: Diagnostic and Prognostic Performance Using Stricter Anatomic CAD Threshold (≥75%/LMCA≥50%)**

#### A. Cohort Characteristics

| Metric | CAD ≥50% | CAD ≥75%/LMCA≥50% | Difference |
| --- | --- | --- | --- |
| Patients with obstructive CAD | 1,357 (60.3%) | 766 (34.0%) | 591 (26.3%) |
| Cohort overlap | — | — | Only 5.4%* |

#### B. Diagnostic Performance

##### hsTnT for Detection of Obstructive Coronary Artery Disease

| Metric | CAD ≥50% | CAD ≥75%/LMCA≥50% | Difference |
| --- | --- | --- | --- |
| Sample size | 2,251 | 2,209 | -42 |
| AUC | 0.676 | 0.668 | -0.008 (p=0.715) |
| Optimal cutoff (Youden) | 6.97 ng/L | 6.97 ng/L | Identical* |
| Sensitivity | 67.1% | 73.2% | +6.1% |
| Specificity | 59.7% | 52.7% | -7.0% |
| PPV | 71.7% | 45.1% | -26.6% |
| NPV | 54.4% | 78.8% | +24.4% |

Incremental diagnostic value of hsTnT (7.0 ng/L cutoff) stratified by RF-CL pre-test probability category is shown below for both obstructive CAD definitions. The inverse gradient of reclassification benefit is fully preserved under the stricter stenosis threshold.

##### RF-CL category-specific diagnostic performance — CAD ≥50% (primary analysis)

| RF-CL Category | n (events) | Prevalence | AUC | Sensitivity | Specificity | NPV | NRI (%) |
| --- | --- | --- | --- | --- | --- | --- | --- |
| Very low | 512 (189) | 36.9% | 0.680 | 70.9% | 57.0% | 77.0% | 38.4 |
| Low | 487 (106) | 21.8% | 0.689 | 56.6% | 71.7% | 85.6% | 19.3 |
| Intermediate/High | 1,252 (593) | 47.4% | 0.622 | 75.0% | 42.5% | 65.4% | 12.4 |
| Overall | **2,251 (888)** | **39.4%** | **0.669** | **72.0%** | **54.1%** | **74.7%** | **—** |

*NRI: net reclassification improvement (reference: ESC 2024 RF-CL model; hsTnT ≥7.0 ng/L as binary classifier). Inverse gradient confirms greatest incremental benefit in lower pre-test probability categories.*

##### RF-CL category-specific diagnostic performance — CAD ≥75%/LMCA≥50% (sensitivity analysis)

| RF-CL Category | n (events) | Prevalence | AUC | Sensitivity | Specificity | NPV | NRI (%) |
| --- | --- | --- | --- | --- | --- | --- | --- |
| Very low | 500 (151) | 30.2% | 0.674 | 72.8% | 55.0% | 82.4% | ≈38.7 |
| Low | 481 (91) | 18.9% | 0.704 | 58.2% | 71.0% | 87.9% | ≈19.1 |
| Intermediate/High | 1,228 (524) | 42.7% | 0.622 | 75.2% | 41.6% | 69.3% | ≈12.9 |
| Overall | **2,209 (766)** | **34.7%** | **0.668** | **72.7%** | **52.8%** | **78.5%** | **—** |

*≈ Estimated from categorical NRI replication; confirm by rerunning original R NRI script with outcome CAD75_HSS50_01. Gradient pattern closely mirrors primary analysis, confirming robustness across stenosis severity definitions.*

#### C. Prognostic Performance

##### NT-proBNP for All-Cause Mortality Prediction in Obstructive CAD

| Metric | CAD ≥50% | CAD ≥75%/LMCA≥50% | Difference |
| --- | --- | --- | --- |
| Sample size | 1,355 | 764 | -591 |
| Deaths | 491 (36.2%) | 307 (40.2%) | +4.0% |
| Median follow-up | 12.6 years | 12.6 years | — |
| NT-proBNP-mortality correlation | r=0.355, p<0.001 | r=0.412, p<0.001 | +0.057 |

#### Key Findings

- **Cohort Overlap:** Only 5.4 percentage point difference demonstrates substantial overlap; results not driven by marginal stenoses.
- **Diagnostic Robustness:** AUC stable (Δ=-0.008, p=0.715) with identical optimal cutoff (6.97 ng/L), confirming threshold robust across severity spectrum.
- **Prognostic Robustness:** Highly significant NT-proBNP-mortality correlations (p<0.001) in both cohorts.

***Abbreviations:*** *AUC = area under receiver operating characteristic curve; CAD = coronary artery disease; hsTnT = high-sensitivity cardiac troponin T; LMCA = left main coronary artery; NPV = negative predictive value; NT-proBNP = N-terminal pro-B-type natriuretic peptide; PPV = positive predictive value.*

**Statistical Methods:** AUC comparison by DeLong test. NT-proBNP-mortality correlation = point-biserial correlation with log-transformed biomarker. Optimal cutoff determined by Youden index (maximizing sensitivity + specificity - 1).

**Supplementary Table S6**

**Missing Data Analysis**

Summary of missing data for all variables used in the primary analyses. The dataset demonstrates excellent completeness with minimal missing data. All biomarkers (NT-proBNP, hsTnT, hsCRP, IL-6, copeptin) have ≥99% complete data. All propensity score matching covariates have ≥95.9% complete data. The low rate of missing data (<5% for all variables) minimizes risk of bias and supports validity of analyses without requiring imputation.

*Abbreviations: NT-proBNP, N-terminal pro-B-type natriuretic peptide; high-sensitivity troponin T, hsTnT; high-sensitivity C-reactive protein, hsCRP; interleukin-6, IL-6; LDL, low-density lipoprotein; HDL, high-density lipoprotein; eGFR, estimated glomerular filtration rate.*

| Category | Variable | N Available | N Missing | % Missing |
| --- | --- | --- | --- | --- |
| Demographics | Age (years) | 2251 | 0 | 0.0% |
|  | Sex (male=1) | 2251 | 0 | 0.0% |
|  | Body mass index (kg/m²) | 2251 | 0 | 0.0% |
|  | Waist-hip ratio | 2158 | 93 | 4.1% |
| Cardiovascular Risk Factors | Smoking status | 2251 | 0 | 0.0% |
|  | Diabetes mellitus | 2251 | 0 | 0.0% |
|  | Hyperlipidemia | 2251 | 0 | 0.0% |
|  | Systolic blood pressure (mmHg) | 2251 | 0 | 0.0% |
|  | Diastolic blood pressure (mmHg) | 2251 | 0 | 0.0% |
| Medications | Statin therapy | 2251 | 0 | 0.0% |
|  | Antihypertensive therapy | 2251 | 0 | 0.0% |
| Clinical History | Family history MI/revascularization <60 years | 2251 | 0 | 0.0% |
| Laboratory Values | eGFR (CKD-EPI, mL/min/1.73m²) | 2251 | 0 | 0.0% |
|  | LDL cholesterol (mmol/L) | 2251 | 0 | 0.0% |
|  | HDL cholesterol (mmol/L) | 2251 | 0 | 0.0% |
|  | Triglycerides (mmol/L) | 2251 | 0 | 0.0% |
| Biomarkers | High-sensitivity CRP (mg/L) | 2228 | 23 | 1.0% |
|  | High-sensitivity troponin T (pg/mL) | 2251 | 0 | 0.0% |
|  | NT-proBNP (pg/mL) | 2251 | 0 | 0.0% |
|  | Interleukin-6 (pg/mL) | 2251 | 0 | 0.0% |
|  | Copeptin (pmol/L) | 2250 | 1 | 0.0% |
| Cardiac Parameters | Left ventricular ejection fraction (%) | 2217 | 34 | 1.5% |
|  | Gensini score | 2248 | 3 | 0.1% |
|  | Number of major epicardial coronary vessels with ≥ one stenosis ≥50% | 2209 | 42 | 1.9% |
| Treatment and Outcomes | Coronary artery disease status | 2251 | 0 | 0.0% |
|  | Revascularization (yes/no) | 2251 | 0 | 0.0% |
|  | All-cause mortality | 2248 | 3 | 0.1% |
|  | Follow-up time (days) | 2245 | 6 | 0.3% |
| Timing | Time from blood draw to catheterization (minutes) | 2168 | 83 | 3.7% |

**Summary:** Mean missing data: 0.43%. Maximum missing: 4.1% (waist-hip ratio). All key biomarkers and outcome variables have ≥99.7% complete data.

**Supplementary Table S7**

**Propensity Score Matching Covariate Balance**

Covariate balance before and after propensity score matching in patients with obstructive coronary artery disease and NT-proBNP ≥150 pg/mL. Standardized mean differences (SMD) quantify covariate imbalance; |SMD| <0.1 indicates excellent balance, <0.15 good balance. Matching used one-to-one nearest-neighbor without replacement (caliper 0.2 SD of logit propensity score) with 19 confounders. Pre-matching mean |SMD| = 0.226 decreased to post-matching mean |SMD| = 0.069, with all covariates achieving |SMD| <0.15. Critical confounders (Gensini score, stenosis count) achieved excellent balance (both |SMD| = 0.028 post-matching). Matching created 572 pairs (1,144 patients total).

*Abbreviations: BMI, body mass index; WHR, waist-hip reatio; BP, blood pressure; CAD, coronary artery disease; eGFR, estimated glomerular filtration rate; HLP, hyperlipoproteinaemia; HDL, high-density lipoprotein; LDL, low-density lipoprotein; LVEF, left ventricular ejection fraction; NT-proBNP, N-terminal pro-B-type natriuretic peptide; PSM, propensity score matching; SD, standard deviation; SMD, standardized mean difference.*

| Covariate | Revasc Mean±SD | No Revasc Mean±SD | Pre-Match SMD | Pre-Match \|SMD\| | Post-Match SMD | Post-Match \|SMD\| |
| --- | --- | --- | --- | --- | --- | --- |
| Age | 66.00±8.82 | 66.55±8.93 | -0.062 | 0.062 | -0.057 | 0.057 |
| Sex | 126.30±44.09 | 131.20±46.39 | -0.108 | 0.108 | -0.142 | 0.142 |
| BMI | 29.69±4.73 | 30.10±4.97 | -0.084 | 0.084 | 0.110 | 0.110 |
| WHR | 0.99±0.08 | 0.98±0.08 | 0.154 | 0.154 | 0.090 | 0.090 |
| SMOKING STATUS | 0.87±0.72 | 0.80±0.74 | 0.090 | 0.090 | 0.024 | 0.024 |
| DIABETEL MELLITUS | 0.39±0.49 | 0.35±0.48 | 0.080 | 0.080 | 0.024 | 0.024 |
| STATIN USE | 0.51±0.50 | 0.45±0.50 | 0.125 | 0.125 | 0.010 | 0.010 |
| ANTIHYPERTENSIVE USE | 0.89±0.32 | 0.93±0.26 | -0.132 | 0.132 | -0.130 | 0.130 |
| FAMILY HISTORY OF PREMATURE CAD | 0.14±0.35 | 0.11±0.32 | 0.069 | 0.069 | 0.090 | 0.090 |
| eGFR_ | 78.24±19.20 | 78.95±19.05 | -0.037 | 0.037 | 0.106 | 0.106 |
| LDL CHOLESTEROL_ | 3.35±1.17 | 3.27±1.10 | 0.072 | 0.072 | -0.004 | 0.004 |
| HDL CHOLESTEROL | 1.28±0.37 | 1.29±0.38 | -0.041 | 0.041 | 0.145 | 0.145 |
| TRIGLYCERIDES | 2.07±2.17 | 2.04±1.36 | 0.017 | 0.017 | -0.125 | 0.125 |
| LVEF | 55.75±11.89 | 55.86±12.00 | -0.009 | 0.009 | 0.033 | 0.033 |
| GENSINI SCORE | 47.58±32.64 | 12.87±20.35 | 1.276 | 1.276 | -0.028 | 0.028 |
| STENOTIC VESSEL COUNT | 2.56±1.45 | 0.45±0.92 | 1.741 | 1.741 | 0.028 | 0.028 |
| HISTORY HLP | 0.90±0.30 | 0.91±0.29 | -0.035 | 0.035 | 0.046 | 0.046 |
| SYSTOLIC BLOOD PRESSURE | 140.70±21.29 | 141.59±19.80 | -0.043 | 0.043 | -0.065 | 0.065 |
| DIASTOLIC BLOOD PRESSURE | 81.01±11.24 | 83.32±11.40 | -0.203 | 0.203 | -0.044 | 0.044 |

**Quality Assessment:** Excellent balance achieved (mean |SMD| = 0.069). Pre-matching imbalances (mean |SMD| = 0.226, maximum 1.741) substantially reduced. All 19 covariates achieved acceptable balance (|SMD| <0.15), meeting published standards (Austin 2011, Stuart 2010).

**Supplementary Table S8A**

**Covariates Retained After Backwards Stepwise Likelihood-Ratio Elimination in Logistic and Cox Regression Models**

Covariate selection used backwards stepwise likelihood-ratio elimination (BSTEP[LR]) with a removal threshold of p>0.10. For each of the five biomarkers, two models were fitted: logistic regression (binary outcome: obstructive CAD ≥50% stenosis at invasive coronary angiography) and Cox proportional hazards regression (outcome: all-cause mortality; events n=644; median follow-up 12.6 years). Covariates marked ✓ were retained in the final model; covariates marked – were eliminated. N/A denotes covariates not included in the candidate pool for that model type.

Abbreviations: BMI, body mass index; CAD, coronary artery disease; eGFR, estimated glomerular filtration rate; HDL, high-density lipoprotein; hs-CRP, high-sensitivity C-reactive protein; hsTnT, high-sensitivity troponin T; IL-6, interleukin-6; LDL, low-density lipoprotein; LVEF, left-ventricular ejection fraction; NT-proBNP, N-terminal pro-B-type natriuretic peptide; N/A, not applicable (covariate not eligible for logistic models). ᵃ Logistic candidate pool (17 covariates): Sex, Age, Diabetes, Smoking, WHR, BMI, Antihypertensives, Systolic BP, Diastolic BP, LDL-C, HDL-C, Triglycerides, eGFR, Statin therapy, Family history of MI <60y, EF, Hyperlipidaemia. Gensini score and number of diseased vessels intentionally excluded from diagnostic models. ᵇ Cox candidate pool (19 covariates): same 17 + Gensini score + number of diseased vessels, entered via a second BSTEP block.7

| **Covariate** | **hsTnT** | | **NT-proBNP** | | **Copeptin** | | **IL-6** | | **hs-CRP** | |
| --- | --- | --- | --- | --- | --- | --- | --- | --- | --- | --- |
|  | **Logit** | **Cox** | **Logit** | **Cox** | **Logit** | **Cox** | **Logit** | **Cox** | **Logit** | **Cox** |
|  | **N=2,135** | **N=2,127** | **N=1,929** | **N=1,893** | **N=1,907** | **N=1,861** | **N=2,129** | **N=2,116** | **N=2,129** | **N=2,116** |
| **Candidate pool (covariates eligible for selection):** | **17ᵃ** | **19ᵇ** | **17ᵃ** | **19ᵇ** | **17ᵃ** | **19ᵇ** | **17ᵃ** | **19ᵇ** | **17ᵃ** | **19ᵇ** |
| **Retained in all applicable logistic AND Cox models** | | | | | | | | | | |
| **Sex** | **✓** | **✓** | **✓** | **✓** | **✓** | **✓** | **✓** | **✓** | **✓** | **✓** |
| **Age** | **✓** | **✓** | **✓** | **✓** | **✓** | **✓** | **✓** | **✓** | **✓** | **✓** |
| **Diabetes mellitus** | **✓** | **✓** | **✓** | **✓** | **✓** | **✓** | **✓** | **✓** | **✓** | **✓** |
| **Smoking status** | **✓** | **✓** | **✓** | **✓** | **✓** | **✓** | **✓** | **✓** | **✓** | **✓** |
| **Body mass index (BMI)** | **✓** | **✓** | **✓** | **✓** | **✓** | **✓** | **✓** | **✓** | **✓** | **✓** |
| **Systolic blood pressure** | **✓** | **✓** | **✓** | **✓** | **✓** | **✓** | **✓** | **✓** | **✓** | **✓** |
| **Diastolic blood pressure** | **✓** | **✓** | **✓** | **✓** | **✓** | **✓** | **✓** | **✓** | **✓** | **✓** |
| **Variably retained — present in ≥1 but not all models** | | | | | | | | | | |
| **Ejection fraction (LVEF)** | **✓** | **✓** | **✓** | **✓** | – | **✓** | – | **✓** | – | **✓** |
| **Waist-hip ratio** | **✓** | – | **✓** | – | **✓** | – | **✓** | – | **✓** | **✓** |
| **LDL cholesterol** | **✓** | – | **✓** | – | **✓** | – | **✓** | – | **✓** | – |
| **HDL cholesterol** | **✓** | **✓** | **✓** | **✓** | **✓** | – | **✓** | **✓** | **✓** | – |
| **Statin therapy** | **✓** | – | **✓** | – | **✓** | – | **✓** | – | **✓** | – |
| **Family history MI <60y** | **✓** | – | **✓** | – | – | – | **✓** | – | **✓** | – |
| **eGFR (CKD-EPI)** | – | **✓** | – | – | – | – | – | **✓** | – | **✓** |
| **Hyperlipidaemia diagnosis** | – | – | – | **✓** | – | **✓** | – | – | – | – |
| **Gensini score** | N/A | **✓** | N/A | **✓** | N/A | **✓** | N/A | **✓** | N/A | **✓** |
| **No. diseased vessels** | N/A | – | N/A | – | N/A | – | N/A | **✓** | N/A | – |
| **Never retained — eliminated in all applicable models** | | | | | | | | | | |
| **Antihypertensives** | – | – | – | – | – | – | – | – | – | – |
| **Triglycerides** | – | – | – | – | – | – | – | – | – | – |
| **Total covariates retained (excluding biomarker):** | **13** | **11** | **13** | **11** | **11** | **10** | **12** | **12** | **12** | **11** |

**Supplementary Table S8B**

**Sensitivity Analysis: Backwards Stepwise Elimination (BSTEP-LR) vs. Forced-Entry (ENTER) — Comparison of Biomarker Effect Estimates**

To assess the influence of covariate selection on biomarker effect estimates, sensitivity analyses were performed forcing all candidate covariates into the model without selection (ENTER method) for all ten models (5 biomarkers × logistic + Cox). OR/HR values represent effect per 1-unit increase in biomarker concentration. Green shading indicates statistical significance (p<0.05); orange shading indicates non-significance. Complete covariate lists for each model are provided in Supplementary Table S9A.

Abbreviations: BSTEP(LR), backwards stepwise likelihood-ratio elimination (removal threshold p>0.10); CI, confidence interval; ENTER, forced-entry method (all candidate covariates simultaneously included); HR, hazard ratio; hs-CRP, high-sensitivity C-reactive protein; hsTnT, high-sensitivity troponin T; IL-6, interleukin-6; NT-proBNP, N-terminal pro-B-type natriuretic peptide; OR, odds ratio. ᵃ n = number of adjustment covariates in final BSTEP model (biomarker not counted).

| **Biomarker** | **Regression model** | **N** | **Covariates** **retained** **(n)ᵃ** | **Backwards elimination**  **OR / HR (95% CI)** | **p** | **Forced-entry ENTER** **OR / HR (95% CI)** | **p** |
| --- | --- | --- | --- | --- | --- | --- | --- |
| **hsTnT** **(per ng/L)** | Logistic regression (CAD ≥50%) | 2,135 | 12 | 1.041 (1.028–1.054) | **<0.001** | 1.042 (1.028–1.056) | **<0.001** |
|  | Cox regression (all-cause mortality) | 2,127 | 11 | 1.025 (1.017–1.034) | **<0.001** | 1.024 (1.015–1.033) | **<0.001** |
| **NT-proBNP** **(per 1 pg/mL)** | Logistic regression (CAD ≥50%) | 1,929 | 13 | 1.002 (1.001–1.002) | **<0.001** | 1.002 (1.001–1.002) | **<0.001** |
|  | Cox regression (all-cause mortality) | 1,893 | 11 | 1.001 (1.001–1.002) | **<0.001** | 1.001 (1.001–1.002) | **<0.001** |
| **Copeptin** **(per 1 pmol/L)** | Logistic regression (CAD ≥50%) | 1,907 | 11 | 1.021 (0.984–1.059) | 0.278 | 1.017 (0.979–1.056) | 0.386 |
|  | Cox regression (all-cause mortality) | 1,861 | 10 | 1.032 (1.001–1.063) | **0.045** | 1.034 (1.002–1.068) | **0.035** |
| **IL-6** **(per 1 pg/mL)** | Logistic regression (CAD ≥50%) | 2,129 | 12 | 1.041 (1.016–1.066) | **0.001** | 1.038 (1.013–1.064) | **0.003** |
|  | Cox regression (all-cause mortality) | 2,116 | 12 | 1.036 (1.023–1.050) | **<0.001** | 1.035 (1.022–1.049) | **<0.001** |
| **hs-CRP** **(per 1 mg/L)** | Logistic regression (CAD ≥50%) | 2,129 | 12 | 1.047 (0.995–1.102) | 0.077 | 1.043 (0.991–1.098) | 0.110 |
|  | Cox regression (all-cause mortality) | 2,116 | 11 | 1.060 (1.020–1.102) | **0.003** | 1.059 (1.018–1.101) | **0.004** |

**STROBE Checklist for Cohort Studies**

Strengthening the Reporting of Observational Studies in Epidemiology (STROBE) statement—checklist of items that should be included in reports of cohort studies.

| **Item** | **Section/Topic** | **Recommendation** | **Page/Line** |
| --- | --- | --- | --- |
| **Title and Abstract** | | | |
| 1 | Title | Indicate the study's design with a commonly used term in the title or abstract | 1 |
| 2 | Abstract | Provide an informative and balanced summary of what was done and what was found | 3-4 |
| **Introduction** | | | |
| 3 | Background/rationale | Explain the scientific background and rationale for the investigation being reported | 6 |
| 4 | Objectives | State specific objectives, including any prespecified hypotheses | 7 |
| **Methods** | | | |
| 5 | Study design | Present key elements of study design early in the paper | 7 |
| 6 | Setting | Describe the setting, locations, and relevant dates, including periods of recruitment, exposure, follow-up, and data collection | 7 |
| 7 | Participants | Give eligibility criteria, and sources and methods of selection of participants. Describe methods of follow-up | 8-9 |
| 8 | Variables | Clearly define all outcomes, exposures, predictors, potential confounders, and effect modifiers | 8-11 |
| 9 | Data sources/measurement | For each variable of interest, give sources of data and details of methods of assessment | 8-11 |
| 10 | Bias | Describe any efforts to address potential sources of bias | 11-14 |
| 11 | Study size | Explain how the study size was arrived at | 7 |
| 12 | Quantitative variables | Explain how quantitative variables were handled in the analyses. If applicable, describe which groupings were chosen and why | 8-11 |
| 13 | Statistical methods | Describe all statistical methods, including those used to control for confounding; methods used to examine subgroups and interactions; how missing data were addressed; sensitivity analyses | 11-14 |
| **Results** | | | |
| 14 | Participants | Report numbers of individuals at each stage of study—e.g., numbers potentially eligible, examined for eligibility, confirmed eligible, included in the study, completing follow-up, and analyzed; give reasons for non-participation at each stage | 15 |
| 15 | Descriptive data | Give characteristics of study participants and information on exposures and potential confounders; indicate number of participants with missing data for each variable of interest; summarize follow-up time | 15, Table 1, Suppl. Table S5 |
| 16 | Outcome data | Report numbers of outcome events or summary measures over time | 19, Figure 3 |
| 17 | Main results | Give unadjusted estimates and, if applicable, confounder-adjusted estimates and their precision (e.g., 95% confidence interval); make clear which confounders were adjusted for and why they were included | 16-19, Figure 1-4, Tables 2 |
| 18 | Other analyses | Report other analyses done—e.g., analyses of subgroups and interactions, and sensitivity analyses | 20, Suppl. Tables S1-S9A/B, Supppl. Figures S1-4 |
| **Discussion** | | | |
| 19 | Key results | Summarize key results with reference to study objectives | 21 |
| 20 | Limitations | Discuss limitations of the study, taking into account sources of potential bias or imprecision. Discuss both direction and magnitude of any potential bias | 24-25 |
| 21 | Interpretation | Give a cautious overall interpretation of results considering objectives, limitations, multiplicity of analyses, results from similar studies, and other relevant evidence | 21-24 |
| 22 | Generalizability | Discuss the generalizability (external validity) of the study results | 25-26 |
| **Other Information** | | | |
| 23 | Funding | Give the source of funding and the role of the funders for the present study and, if applicable, for the original study on which the present article is based | 26 |

*Note: Page numbers refer to the main manuscript unless otherwise specified (e.g., "Suppl." for supplementary materials).*
